# Occupational differences in COVID-19 hospital admission and mortality risks between women and men in Scotland: a population-based study using linked administrative data

**DOI:** 10.1101/2024.01.25.24301783

**Authors:** Serena Pattaro, Nick Bailey, Chris Dibben

## Abstract

**Introduction:** Occupations vary with respect to workplace factors that influence exposure to COVID-19, such as ventilation, social contacts and protective equipment. Variations between women and men may arise because they have different occupational roles or behavioural responses. We estimate occupational differences in COVID-19 hospital admission and mortality risks by sex.

**Methods:** We combined individual-level data from 2011 Census with (i) health records and (ii) household-level information from residential identifiers. We used data for a cohort of 1.7 million Scottish adults aged 40-64 years between 1 March 2020 and 31 January 2021. We estimated age-standardised COVID-19 hospital admission and mortality rates, stratified by sex and occupation. Using Cox proportional hazards models, we estimated COVID-19 hospital admission and death risks, adjusting for relevant factors.

**Results:** Generally, women had lower age-standardised COVID-19 hospital admission and mortality rates compared to men. Among women, adjusted death risks were lower for health professionals, and those in associate professional and technical occupations (paramedics and medical technicians). Among men, elevated adjusted admission and death risks were observed for large vehicle and taxi drivers. Additionally, admission risks remained high among men working in caring personal services, including home and care workers, while elevated risks were observed among women working in customer service occupations (call centre operators) and as process, plant and machine operatives (assemblers/sorters).

**Conclusion:** Occupational differences in COVID-19 hospital admission and mortality risks between women and men highlight the need to account for sex differences when developing interventions to reduce infections among vulnerable occupational groups.

## Introduction

The COVID-19 pandemic had starkly unequal effects on working-age women and men, with evidence suggesting higher rates of severe COVID-19 outcomes, including hospitalisation and mortality, among men [1]. While infection rates were elevated among women of working age compared to those of younger and retirement ages [2, 3], women, in contrast to men, appeared relatively protected from more adverse health consequences, potentially due to differing biological responses. Disparities in severe COVID-19 outcomes have been linked to different biological markers underpinning sex-based immune responses to viral infection [4, 5].

Beyond biological factors, socio-economic and behavioural characteristics likely influence the unequal distribution of severe COVID-19 outcomes between women and men, with occupation and workplace settings playing a crucial role in shaping exposure and transmission [6]. Key occupational exposure factors include the frequency of contacts with other people, the physical proximity and ability to maintain social distance, the physical work environment (whether indoors or outdoors), and the availability of appropriate protective equipment [7]. According to one US study, women tend to be employed in more contact-intensive occupations with a higher degree of physical proximity to others [8]. These roles are concentrated in the service sector, including health and social care, food preparation and education, where essential and/or frontline occupations are over represented [9]. Men, on the other hand, tend to be employed in production occupations in the construction and manufacturing sectors with a lower degree of contact intensity [8]. A recent study of England and Wales found that workplace contacts partially accounted for the increased COVID-19 infection risks for workers in healthcare professions, as well as indoor trade, process and plant occupations. These occupational groups also had a higher risk of working in poorly ventilated workplaces, highlighting the role played by occupational factors in viral exposure [10].

Some occupational groups may not only be at increased risk of exposure, but also increased risk of adverse outcomes. Population-based studies from the UK have found higher COVID-19 mortality risks among male taxi and cab drivers, and among female health and social care professional occupations [11, 12] although, in the latter group, the risks were attenuated after the first pandemic wave [13]. A similar temporal pattern was reported by a case-control study in Scotland which found a higher risk of both hospital admission and severe COVID-19, including mortality, among healthcare workers and their household’s members [14].

Behavioural factors are potentially another important driver of occupational and gender disparities in COVID-19 outcomes. Factors may include risk-taking behaviour, compliance with preventive measures (e.g. hygiene practices, mask wearing and use of protective equipment), and seeking/receiving health care. An international study conducted during the first pandemic wave found that women were more likely to perceive COVID-19 as a serious health problem and to comply with government-imposed restriction measures [15]. Conversely, men are generally more likely to engage in high-risk and health-damaging behaviours, including smoking and excessive alcohol consumption [16]. Men are also less likely to seek/receive health care, potentially resulting in delays in treatment for severe disease and leading to more severe outcomes [17].

Occupational disparities in COVID-19 outcomes are also associated with household circumstances and pre-pandemic health conditions. People with lower-paid, lower-skilled and more precarious jobs are more likely to live in overcrowded accommodations and co-reside with younger adults or children, increasing the exposure risks to viral transmission. They are also more likely to have comorbid conditions, including cardiovascular disease, diabetes, cancer, respiratory problems, liver disease, and renal disease [17]. Many of these conditions have been associated with increased risks of COVID-19 mortality in England [18]. Another English study found that living in a multigenerational household was associated with an elevated risk of COVID-19 mortality among older adults from ethnic minority groups, including women [19].

Occupational differences in COVID-19 outcomes for working-age women and men are likely driven by occupational and wider socio-economic, including household factors. It is important to understand the relative contribution of each to inform future interventions. In this study, we leveraged a novel Scottish data collection, described in the “Data and study population” section. We aimed to investigate how occupational differences in COVID-19 admissions and mortality differ across women and men between 1 March 2020 and 31 January 2021, using Cox proportional hazards models and adjusting for socio-economic and pre-pandemic health factors.

## Methods

### Data and study population

We used a large data collection covering the population of Scotland. This combined (i) individual-level demographic and socio-economic information from 2011 Census; (ii) household-level information from Ordnance Survey’s residential identifiers; (iii) occupational exposure measures from Occupational Information Network (O*NET) survey data; and (iv) electronic health information on mortality, hospitalisation, laboratory testing and primary care records from Public Health Scotland’s (PHS) COVID-19 Research Database [20]. The data collection is part of a wider study investigating social risk factors for COVID-19 in Scotland [21], which was established as part of the Scottish Data and Intelligence Network [22] to inform the government’s response to the pandemic.

The Scottish population spine includes all individuals registered with a General Practice, who have received a unique Community Health Index (CHI) number. We restrict our study to those in the 2011 Census with linkable records to the population spine. The Census provides occupation as recorded in 2011, which we use as a measure of likely occupation in 2020/21. We exclude those who died or left Scotland before 1 March 2020 (date of the first confirmed positive COVID-19 case in Scotland). The population spine (CHI register) provides the latest address notified to the General Practice. The address text was used to attach a household identifier, derived from Ordnance Survey’s Unique Property Reference Numbers (UPRNs), through an innovative residential linkage tool [23]. Following subsequent linkage to electronic health records from Public Health Scotland’s COVID-19 Research Database, data were available for 4.9 million people (approximately 90% of the Scottish population [24]). The population of interest was those aged 40-64 years on 1 March 2020. From the initial study population, we excluded those aged under 40 or above 64 years, yielding a cohort of approximately 1.7 million individuals (Fig. 1).

**Fig. 1.**
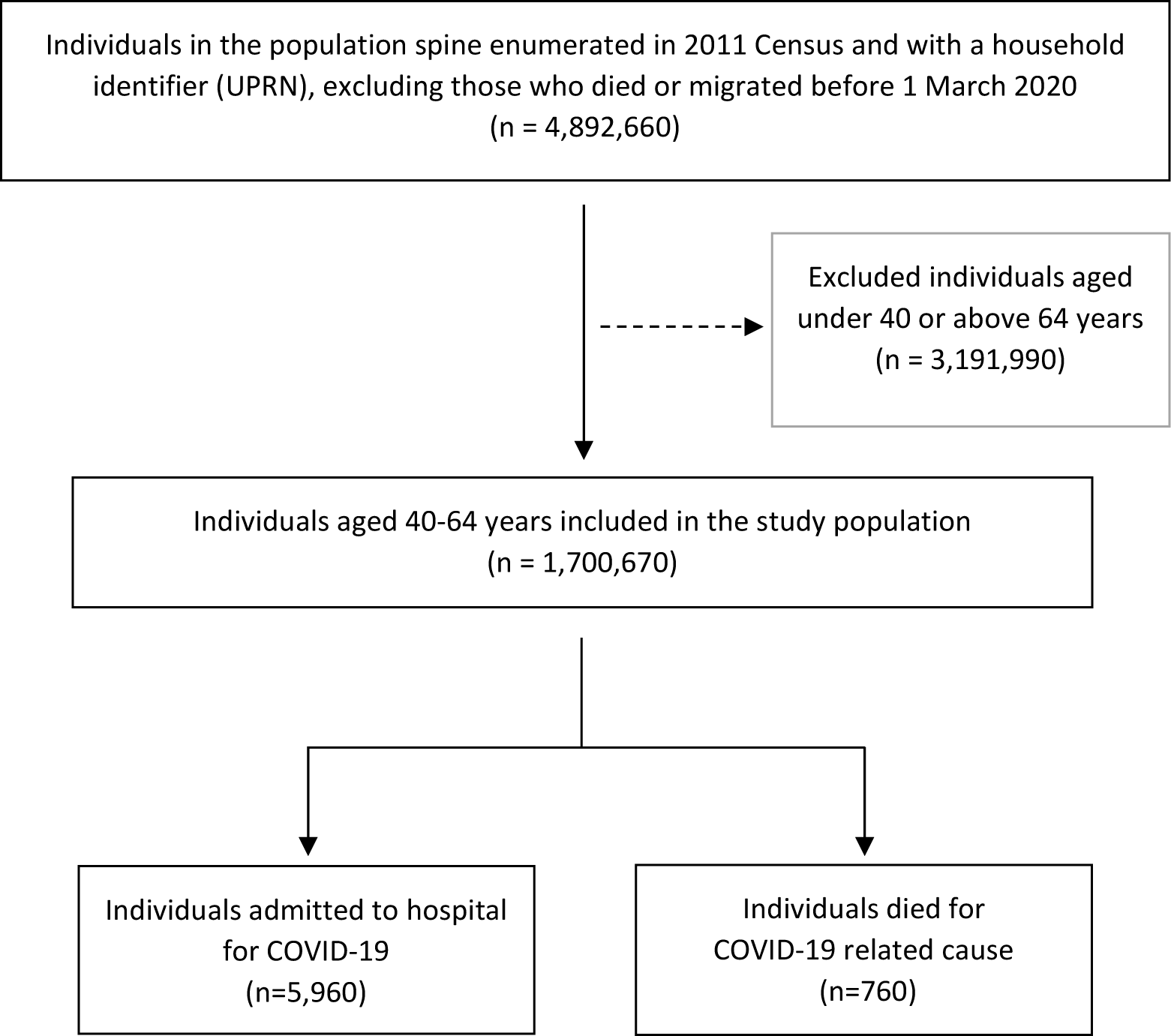
Flow chart representing the selection process for the study population. ^a^ Reported figures are rounded to the nearest 10.

### Outcomes

The outcomes of interest were time to first COVID-19 hospital admission and death. Hospital admission was identified using International Classification of Diseases 10^th^ edition (ICD-10) codes for confirmed (U07.1) or suspected COVID-19 (U07.2) as primary or secondary cause, or an admission occurring within 14 days from a laboratory-confirmed COVID-19 infection. Death was defined using ICD-10 codes for confirmed or suspected COVID-19 (primary or secondary cause), reported on death certificates and identified within 28 days of a laboratory-confirmed COVID-19 infection. These outcome definitions are in line with those used in Scottish official statistics [25].

### Exposure

We constructed a measure of occupation using the UK Standard Occupational Classification (SOC) 2010 reported in 2011 Census [26], capturing the occupational structure and skill levels required to perform a role. We identified 26 occupational groups by combining different levels of the hierarchical classification, ranging from major groups at the highest level (one-digit codes) to more detailed unit groups (four-digit codes) (Supplementary Table 1). The selected occupational groups enabled us to identify specific occupations which were shown to be at high risk of COVID-19 outcomes (taxi drivers, social care and health care workers), while allowing for comparison with previous studies [11-14].

Some discrepancies may arise between the occupational information reported in 2011 Census and actual occupation in March 2020. Note that individuals may have experienced job displacement during the COVID-19 pandemic. Since both add *noise* to our measure, estimates of the relationships between occupation and COVID-19 risks should therefore be regarded as lower limits. Previous studies have shown that women experienced greater job losses than men [8]. Using data from the UK Household Longitudinal Study (UKHLS) [27], we computed the proportion of men and women aged 40-64 years who retained the same occupation between 2011 and 2020 using major group occupations. For women, proportions varied between 40.8% among managers, directors and senior officials and 74.1% among workers in caring, leisure and other service occupations. For men, proportions varied between 38.8% among those employed in sales and customer service occupations and 72% among process, plant and machine operatives (Supplementary Table 2). Despite the limited disaggregation possible with the UKHLS, the descriptive statistics suggests that the occupational information in 2011 Census is relatively stable over time.

### Covariates

Covariates used in the statistical analysis were characteristics that may be associated with both occupation and COVID-19 hospital admission and mortality risks (Supplementary Table 3). Some characteristics may act as either potential confounders or mediators, but it is difficult to separate them out due to data limitations. Socio-demographic characteristics were age and sex derived from the CHI register, and ethnicity taken from 2011 Census. For household-level characteristics, we included housing tenure, reported in 2011 Census, and household size, whether the household included children, and whether the household included multiple generations based on people’s age cohort, constructed from household identifiers based on UPRN. We also included occupational exposure information from three score measures assessing the frequency of exposure to disease/infection, the extent of physical proximity to others, and the frequency of working in environmentally-controlled indoor settings. Score measures were sourced from US survey data from Occupational Information Network (O*NET) [28], mapped onto UK SOC 2010 codes, via ISCO-08 codes, and then standardised using a procedure described by Office for National Statistics [29]. Health-related characteristics included whether individuals had a learning disability/difficulty recorded in 2011 Census, and whether they were shielding (to minimise physical contacts with others) due to underlying vulnerable conditions (PHS shielding patient list dataset) (Supplementary Table 4). Pre-pandemic health conditions were derived from cluster variables based on Read codes recorded in primary care data from PHS COVID-19 Research Database. Cancer and immunosuppression, cardiovascular conditions, diabetes, hypertension, respiratory conditions, and other conditions were considered factors likely to heighten severe COVID-19 risks based on existing studies [18, 30] (Supplementary Table 5).

### Statistical analysis

We summarised the individual- and household-level characteristics for the total population and for those who were first admitted to hospital and died for COVID-19. We reported baseline characteristics for the covariates of interest using counts and percentages. We also calculated annualised age-standardised COVID-19 hospital admission and mortality rates per 100,000 persons for each occupational group. Rates were estimated separately for women and men, using a direct method and standardised to the 2013 European Standard Population [31]. Confidence intervals were calculated using the method proposed by Dobson et al. [32] accounting for the small number of observed events. Estimates were produced using ‘distrate’ command [33, 34] in Stata/MP 16 (StataCorp LP, College Station, Texas).

We modelled the time to COVID-19 hospital admission and death, separately for women and men, using Cox proportional hazards models. Hazard ratios were interpreted as the rate at which an event of interest occurs in one group relative to the rate at which it occurs in a reference group over time. We estimated a set of nested models to adjust for potential confounders. First, we adjusted for individual-level socio-demographic factors including age and ethnicity. We used a restricted cubic spline to model the non-linear association between age and COVID-19 outcomes. We then included household-level characteristics and measures of occupational exposure that are likely to confound the relationship between workplace factors and COVID-19 admission and death. In the last model, we further adjusted for confounders such as disability and health-related factors including whether the person had a learning disability/difficulty or whether shielding, and pre-pandemic health conditions. Model fit was assessed using Akaike Information Criterion (AIC) and Bayesian Information Criterion (BIC) statistics. Analyses were conducted in the Scottish National Safe Haven using Stata/MP 16 and R (version 3.6.1).

## Results

The study population covered 1,700,670 Scottish adults aged 40-64 years, who were observed between 1 March 2020 and 31 January 2021. The mean age was 52.4 years (standard deviation (SD) 7.0) and 50.6% of adults were women. Individuals were followed up for 1,514,189 person-years until first COVID-19 admission to hospital, 1,561,010 person-years until COVID-19 death, or the end of follow-up period on 31 January 2021. Among 5,960 COVID-19 admissions, the median follow-up time was 232 days (95% confidence interval (CI) 228-233); COVID-19 admissions were more likely to occur among men (52.7%) and older individuals (mean age 54.7 years (SD 6.4)). Among the 760 COVID-19 deaths, the median follow-up time was 248 days (95%CI 237-253), with 460 deaths (60.5%) occurring among men and a mean age of 57.2 years (SD 5.8). Additional baseline characteristics of the population, overall and by outcome of interest are described in Table 1.

**Table 1.** Characteristics of the study population and those who were admitted to hospital or died for a COVID-19 cause.

| Characteristics | Population <sup>a</sup> |  | COVID-19 hospital admissions <sup>a</sup> |  | COVID-19 deaths <sup>a</sup> |  |
| --- | --- | --- | --- | --- | --- | --- |
|  | (n=1,700,670) |  | (n=5,960) |  | (n=760) |  |
|  | N | % | n | % | n | % |
| *Sex:* |  |  |  |  |  |  |
| Women | 859,800 | 50.56 | 2,820 | 47.32 | 300 | 39.47 |
| Men | 840,870 | 49.44 | 3,140 | 52.68 | 460 | 60.53 |
| *Age(years) <sup>b</sup>* | 52.40 | 7.00 | 54.70 | 6.40 | 57.20 | 5.80 |
| *Ethnicity:* |  |  |  |  |  |  |
| Non-white | 195,810 | 11.51 | 770 | 12.92 | 90 | 11.84 |
| White | 1,504,860 | 88.49 | 5,190 | 87.08 | 670 | 88.16 |
| *Occupation (SOC 2010 codes):* |  |  |  |  |  |  |
| 1 - Managers, directors and senior officials | 136,730 | 8.04 | 460 | 7.72 | 50 | 6.58 |
| 2 - Professional occupations | 118,250 | 6.95 | 190 | 3.19 | 20 | 2.63 |
| 22 - Health professionals | 74,150 | 4.36 | 310 | 5.20 | 10 | 1.32 |
| 24 - Business, media and public service professionals | 65,340 | 3.84 | 120 | 2.01 | 10 | 1.32 |
| 3 - Associate professional and technical occupations | 76,370 | 4.49 | 190 | 3.19 | 20 | 2.63 |
| 33 - Protective service occupations | 26,910 | 1.58 | 100 | 1.68 | 10 | 1.32 |
| 35 - Business and public service associate professionals | 88,630 | 5.21 | 170 | 2.85 | 20 | 2.63 |
| 41 - Administrative occupations | 138,500 | 8.14 | 390 | 6.54 | 50 | 6.58 |
| 42 - Secretarial and related occupations | 40,180 | 2.36 | 120 | 2.01 | 10 | 1.32 |
| 5 - Skilled trades occupations | 90,400 | 5.32 | 290 | 4.87 | 30 | 3.95 |
| 53 - Skilled construction and building trades | 60,580 | 3.56 | 220 | 3.69 | 30 | 3.95 |
| 543 - Food preparation and hospitality trades | 29,060 | 1.71 | 110 | 1.85 | 20 | 2.63 |
| 6 - Caring, leisure and other service occupations | 70,140 | 4.12 | 230 | 3.86 | 20 | 2.63 |
| 614 - Caring personal services | 28,470 | 1.67 | 180 | 3.02 | 10 | 1.32 |
| 6145 - Care workers and home workers | 52,310 | 3.08 | 300 | 5.03 | 30 | 3.95 |
| 71 - Sales occupations | 87,000 | 5.12 | 320 | 5.37 | 50 | 6.58 |
| 72 - Customer service occupations | 26,820 | 1.58 | 120 | 2.01 | 10 | 1.32 |
| 81 - Process, plant and machine operatives | 41,230 | 2.42 | 180 | 3.02 | 30 | 3.95 |
| 811 - Process plant operatives | 19,940 | 1.17 | 90 | 1.51 | 20 | 2.63 |
| 82 - Transport and mobile machine drivers and operatives | 40,830 | 2.40 | 210 | 3.52 | 30 | 3.95 |
| 8211 - Large goods vehicle drivers | 17,040 | 1.00 | 90 | 1.51 | 20 | 2.63 |
| 8214 - Taxi and cab drivers and chauffeurs | 12,170 | 0.72 | 110 | 1.85 | 20 | 2.63 |
| 9 - Elementary occupations | 59,540 | 3.50 | 240 | 4.03 | 40 | 5.26 |
| 91 - Elementary trades and related occupations | 34,600 | 2.03 | 130 | 2.18 | 30 | 3.95 |
| 927 - Other elementary services occupations | 38,440 | 2.26 | 160 | 2.68 | 20 | 2.63 |
| 9233 - Cleaners and domestics | 41,680 | 2.45 | 200 | 3.36 | 30 | 3.95 |
| No code required | 185,360 | 10.90 | 730 | 12.25 | 120 | 15.79 |
| *Housing tenure:* |  |  |  |  |  |  |
| Owned outright | 226,160 | 13.30 | 600 | 10.07 | 80 | 10.53 |
| Owned with mortgage | 897,530 | 52.78 | 2,960 | 49.66 | 300 | 39.47 |
| Social rented | 282,060 | 16.59 | 1,520 | 25.50 | 260 | 34.21 |
| Private rented | 138,700 | 8.16 | 380 | 6.38 | 50 | 6.58 |
| Owned/not known | 156,220 | 9.19 | 500 | 8.39 | 70 | 9.21 |
| *Household size:* |  |  |  |  |  |  |
| 1-2 people | 779,040 | 45.81 | 2,860 | 47.99 | 420 | 55.26 |
| 3-4 people | 720,230 | 42.35 | 2,310 | 38.76 | 220 | 28.95 |
| 5-6 people | 160,170 | 9.42 | 540 | 9.06 | 40 | 5.26 |
| 7+ people | 41,230 | 2.42 | 250 | 4.19 | 80 | 10.53 |
| *Household with children:* |  |  |  |  |  |  |
| No children | 1,300,090 | 76.45 | 4,920 | 82.55 | 690 | 90.79 |
| At least one child 0-11 years | 187,120 | 11.00 | 440 | 7.38 | 30 | 3.95 |
| At least one child 12-17 years | 213,460 | 12.55 | 600 | 10.07 | 40 | 5.26 |
| *Whether multigenerational household:* |  |  |  |  |  |  |
| Yes | 141,970 | 8.35 | 530 | 8.89 | 640 | 84.21 |
| No | 1,558,700 | 91.65 | 5,430 | 91.11 | 120 | 15.79 |
| *Occupational exposure measures <sup>b</sup>:* |  |  |  |  |  |  |
| Exposure to disease | 0.30 | 0.30 | 0.30 | 0.30 | 0.30 | 0.30 |
| Proximity to others | 0.60 | 0.20 | 0.60 | 0.30 | 0.50 | 0.30 |
| Environmentally controlled indoor conditions | 0.70 | 0.30 | 0.60 | 0.30 | 0.60 | 0.30 |
| *Learning disability or difficulty:* |  |  |  |  |  |  |
| Yes | 31,770 | 1.87 | 210 | 3.52 | 50 | 6.58 |
| No | 1,518,930 | 89.31 | 5,310 | 89.09 | 660 | 86.84 |
| Not known | 149,970 | 8.82 | 440 | 7.38 | 50 | 6.58 |
| *Whether shielding:* |  |  |  |  |  |  |
| Yes | 39,100 | 2.30 | 630 | 10.57 | 150 | 19.74 |
| No | 1,661,570 | 97.70 | 5,330 | 89.43 | 610 | 80.26 |
| *Pre-pandemic health conditions* |  |  |  |  |  |  |
| *Cancer and immunosuppression:* |  |  |  |  |  |  |
| Yes | 23,960 | 1.41 | 190 | 3.19 | 30 | 3.95 |
| No | 1,676,710 | 98.59 | 5,770 | 96.81 | 730 | 96.05 |
| *Cardiovascular conditions:* |  |  |  |  |  |  |
| Yes | 118,880 | 6.99 | 960 | 16.11 | 190 | 25.00 |
| No | 1,581,790 | 93.01 | 5,000 | 83.89 | 570 | 75.00 |
| *Diabetes:* |  |  |  |  |  |  |
| Yes | 104,150 | 6.12 | 1,130 | 18.96 | 200 | 26.32 |
| No | 1,596,520 | 93.88 | 4,830 | 81.04 | 560 | 73.68 |
| *Hypertension:* |  |  |  |  |  |  |
| Yes | 241,210 | 14.18 | 1,590 | 26.68 | 230 | 30.26 |
| No | 1,459,460 | 85.82 | 4,370 | 73.32 | 530 | 69.74 |
| *Respiratory conditions:* |  |  |  |  |  |  |
| Yes | 246,490 | 14.49 | 1,420 | 23.83 | 210 | 27.63 |
| No | 1,454,180 | 85.51 | 4,540 | 76.17 | 550 | 72.37 |
| *Other conditions:* |  |  |  |  |  |  |
| Yes | 308,710 | 18.15 | 1,650 | 27.68 | 270 | 35.53 |
| No | 1,391,960 | 81.85 | 4,310 | 72.32 | 490 | 64.47 |

<sup>a</sup> Hospital admissions and deaths occurring between 1 March 2020 and 31 January 2021; reported figures are rounded to the nearest 10.
<sup>b</sup> Mean and standard deviation are reported.

Table 2 shows annualised age-standardised COVID-19 hospital admission and death rates for women and men aged 40-64 years. Generally, women had lower age-standardised rates (ASRs) than men across both outcomes. For women, the highest ASRs for COVID-19 admissions (highlighted in bold) were observed among those working in caring personal services, including nursing assistants and ambulance staff (excluding paramedics), with 599.7 admissions (95%CI 499.0-714.1) per 100,000 persons, and process, plant and machine operatives in the food, drink and tobacco industry (assemblers/sorters), with 576.8 admissions (95%CI 426.9-645.0). For men, the highest ASRs for COVID-19 admissions were observed among taxi and cab drivers, with 949.8 admissions per 100,000 persons (95%CI 763.8-1164.6), followed by workers in caring personal services, with 916.2 admissions (95%CI 663.0-1231.7) per 100,000 persons. For both women and men, the lowest (or second-lowest) rates (highlighted in white, grey cells) were observed among professionals in science, research, engineering, and technology, and in teaching and education, with 150.9 admissions (95%CI 120.4-186.8) among women and 190.4 admissions (95%CI 155.7-230.6) among men.

**Table 2.** Annualised age-standardised rates (ASR) for COVID-19 hospital admissions and deaths per 100,000 persons aged 40-64 years in Scotland, by occupation and sex.

| SOC<br>2010<br>code | Occupation | COVID-19 hospital admissions <sup>a</sup> |  | COVID-19 deaths <sup>a</sup> |  |
| --- | --- | --- | --- | --- | --- |
|  |  | Women<br>(n=2820)<br>ASR (95%CI) <sup>b</sup> | Men<br>(n=3140)<br>ASR (95%CI) <sup>b</sup> | Women<br>(n=300)<br>ASR (95%CI) <sup>b</sup> | Men<br>(n=460)<br>ASR (95%CI) <sup>b</sup> |
| 1 | Managers, directors and senior officials | 255.6 (214.2-302.6) | 382.6 (340.5-428.3) | 29.4 (16.8-47.9) | 40.8 (28.2-57.1) |
| 2 | Professional occupations | **150.9 (120.4-186.8)** | **190.4 (155.7-230.6)** | **15.4 (7.0-29.3)** | **21.1 (10.9-36.8)** |
| 22 | Health professionals | 433.9 (380.8-492.2) | 511.6 (392.8-654.7) | 19.5 (9.7-34.9) | **8.8 (0.2-48.8)** |
| 24 | Business, media and public service professionals | **130.8 (91.0-182.2)** | **238.0 (188.6-296.2)** | - | **28.4 (13.6-52.4)** |
| 3 | Associate professional and technical occupations | 218.6 (172.6-272.9) | 325.0 (268.7-389.5) | **10.5 (2.9-26.9)** | 43.7 (25.0-71.0) |
| 33 | Protective service occupations | - | 457.6 (368.3-561.8) | - | 51.6 (25.4-92.9) |
| 35 | Business and public service associate professionals | **161.9 (123.9-207.9)** | **252.2 (206.9-304.4)** | **7.9 (1.6-23.1)** | 35.9 (20.5-58.4) |
| 41 | Administrative occupations | 277.8 (246.5-311.9) | 371.4 (302.4-451.4) | 30.2 (20.7-42.5) | 62.6 (37.0-99.1) |
| 42 | Secretarial and related occupations | 303.0 (247.8-366.4) | - | 29.5 (14.7-52.4) | - |
| 5 | Skilled trades occupations | 226.1 (127.2-368.6) | 327.1 (288.7-369.2) | 34.8 (1.8-132.8) | 33.0 (22.1-47.4) |
| 53 | Skilled construction and building trades | - | 369.8 (322.0-422.6) | - | 52.4 (35.7-74.1) |
| 543 | Food preparation and hospitality trades | 346.2 (251.9-463.7) | 451.7 (349.8-573.9) | 28.0 (7.6-71.7) | 76.4 (38.0-137.0) |
| 6 | Caring, leisure and other service occupations | 327.7 (281.8-378.9) | 472.9 (345.9-630.5) | 18.8 (8.9-34.7) | 64.5 (22.7-142.4) |
| 614 | Caring personal services | **599.7 (499.0-714.1)** | **916.2 (663.0-1231.7)** | 42.4 (19.1-79.9) | 45.1 (3.9-167.6) |
| 6145 | Care workers and home workers | **566.4 (494.9-645.0)** | **736.1 (553.3-959.2)** | 60.5 (39.6-88.3) | 59.9 (19.5-139.8) |
| 71 | Sales occupations | 365.3 (320.3-414.8) | 477.6 (378.9-593.9) | 50.9 (35.4-70.8) | 84.8 (46.2-142.4) |
| 72 | Customer service occupations | 499.9 (398.6-619.0) | 457.8 (315.9-641.1) | 41.3 (16.6-85.2) | 54.3 (14.7-139.4) |
| 81 | Process, plant and machine operatives | **576.8 (426.9-759.7)** | 392.2 (324.7-469.4) | **108.6 (57.6-186.2)** | 54.4 (31.4-87.6) |
| 811 | Process plant operatives | 440.3 (299.2-622.6) | 411.6 (305.6-542.3) | 40.9 (11.1-104.9) | 84.4 (42.1-151.2) |
| 82 | Transport and mobile machine drivers and operatives | 336.3 (159.5-621.4) | 498.4 (428.8-575.9) | 31.3 (0.8-174.2) | 60.8 (38.8-90.5) |
| 8211 | Large goods vehicle drivers | - | 501.2 (392.5-628.9) | - | **95.7 (56.2-151.9)** |
| 8214 | Taxi and cab drivers and chauffeurs | - | **949.8 (763.8-1164.6)** | - | **132.9 (79.4-208.5)** |
| 9 | Elementary occupations | 310.7 (225.4-416.7) | 441.5 (381.1-508.6) | **77.0 (39.3-135.3)** | 64.2 (42.9-92.3) |
| 91 | Elementary trades and related occupations | 418.8 (282.1-596.9) | 377.2 (306.4-459.3) | **127.6 (55.6-246.7)** | 67.2 (39.7-106.3) |
| 927 | Other elementary services occupations | 403.3 (331.9-485.5) | 605.7 (447.6-801.4) | 33.5 (16.1-61.7) | **149.0 (76.8-260.4)** |
| 9233 | Cleaners and domestics | 452.4 (385.8-526.9) | 342.9 (205.2-537.2) | 56.9 (36.3-85.0) | 74.1 (19.7-190.9) |

<sup>a</sup> Hospital admissions and deaths occurring between 1 March 2020 and 31 January 2021. <sup>b</sup> 95% Confidence Intervals (CI) calculated using the Dobson method [32] are reported; ASRs are standardised to the 2013 European Standard population using the direct method, and annualised; estimates are not reported if there were less than 10 admissions or deaths; for each sex the three highest ASRs are highlighted in bold and the three lowest ASRs are highlighted in white (grey cells).

The ASRs for COVID-19 mortality show a steeper occupational gradient compared to the ASRs for hospital admission. Among women, those working in elementary trades and related occupations (packers and canners) recorded the highest ASR of 127.6 (95%CI 55.6-246.7) COVID-19 deaths per 100,000 persons, while business and public service associate professionals, including those in the transport and legal administration sectors, had the lowest ASRs with 7.9 (95%CI 1.6-23.1) deaths per 100,000 persons. For men, the highest ASR was 149 (95%CI 76.8-260.4) deaths per 100,000 persons among those working in other elementary services occupations (kitchen assistants and waiters), while one of the lowest ASRs of 21.1 (95%CI 10.9-36.8) deaths was recorded among those in professional occupations.

On average the relative difference in the risks of COVID-19 hospital admission between the occupational groups of interest and managers, directors and senior officials (reference category) decreased after adjusting for confounding factors for both women and men (Fig. 2; Supplementary Table 6 for full-model details). However, for some occupational groups, risks remained significantly higher relative to the reference category. For example, in the fully-adjusted model for women, those working in customer service occupations had a hazard ratio (HR) of 1.61 (95%CI 1.22-2.11) compared to 1.93 (95%CI 1.47-2.54) in the baseline model adjusting for age and ethnicity. An elevated risk also remained for process, plant and machine operatives (assemblers/sorters) with HR of 1.49 (95%CI 1.05-2.11). Among men, higher risks were observed among taxi and cab drivers (HR 1.84, 95%CI 1.42-2.40), those working in caring personal services (ambulance staff) (HR 1.57, 95%CI 1.11-2.22), and care and home workers (HR 1.38, 95%CI 1.00-1.91).

**Fig. 2.**
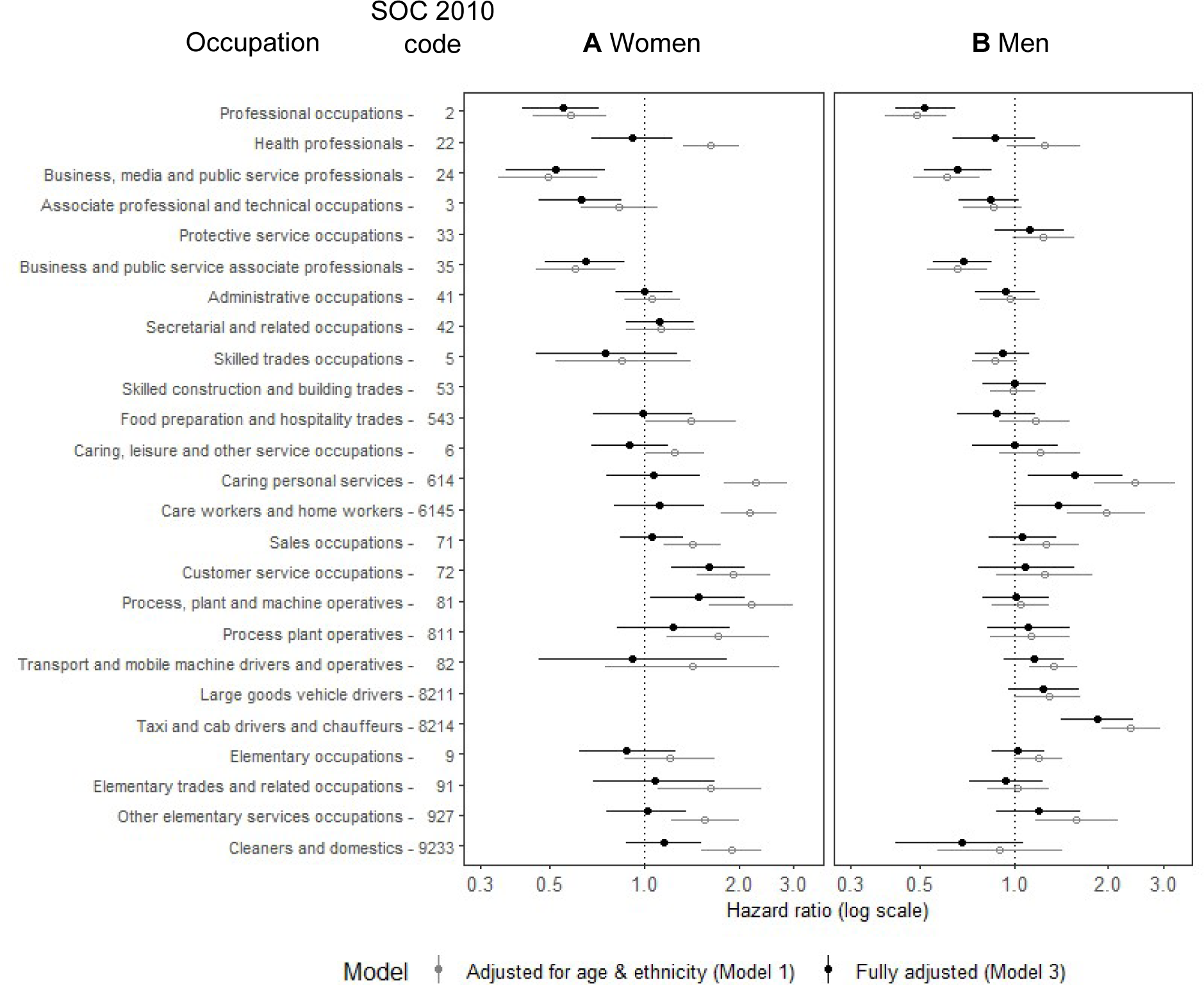
Hazard ratios for COVID-19 hospital admission for (A) women and (B) men aged 40-64 years in Scotland. _a_ Hazard ratios and 95% confidence intervals are displayed on a logarithmic scale; reference category: 1 ‘Managers, directors and senior officials’; fully-adjusted model (Model 3) is coloured in black (solid circle) and appears above Model 1 (grey hollow circle) for each group; fully-adjusted model (Model 3, Supplementary Table 6) additionally controls for: housing tenure, household size, household with children, whether multigenerational household, occupational exposure measures, learning disability or difficulty, whether shielding and pre-pandemic health conditions; hospital admissions occurring between 1 March 2020 and 31 January 2021; data are not reported if there were less than 10 hospital admission events.

For both women and men, lower COVID-19 admission risks were observed among those in higher occupational groups, after adjusting for household characteristics and pre-pandemic health conditions. For example, among professional occupations, women reported an HR of 0.55 (95%CI 0.41-0.72) compared to 0.58 (95%CI 0.44-0.76) in the baseline model. The corresponding HRs for men were 0.52 (95%CI 0.42-0.65) and 0.49 (95%CI 0.39-0.61). Lower risks were also observed among both female and male professionals in business, media and public service (adjusted HR 0.52, 95%CI 0.36-0.75 and 0.66, 95%CI 0.52-0.85, respectively) and associate professionals in business and public service (adjusted HR 0.65, 95%CI 0.48-0.87 and 0.69, 95%CI 0.55-0.85, respectively).

More distinct differences between women and men emerged when considering the risk of COVID-19 death (Fig. 3; see Supplementary Table 7 for full model details). Female health professionals (medical practitioners, nurses and pharmacists) had a lower death risk relative to the reference group, after adjusting for confounders (HR 0.28, 95%CI 0.10-0.78). A lower risk was also observed among women in associate professional and technical occupations, including those working in health and social care, and in science, engineering and technology (HR 0.24, 95%CI 0.07-0.78), and those in caring, leisure and other service occupations (childminders and nursery assistants) (HR 0.36, 95%CI 0.14-0.92). Conversely, among men, after adjusting for socio-economic and health factors, increased COVID-19 death risks were displayed by taxi and cab drivers (HR of 3.46, 95%CI 1.74-6.86) and large goods vehicle drivers (HR 2.69, 95%CI 1.45-4.99). For the latter group, there was an increase in COVID-19 death risk compared to the risk recorded in the baseline model (HR 2.34, 95%CI 1.32-4.13). Elevated risks were also shown for those working in other elementary services occupations, including kitchen assistants and waiters, (adjusted HR 2.44, 95%CI 1.21-4.91) and food, drink and tobacco process plant operatives (adjusted HR 2.39, 95%CI 1.16-4.91).

**Fig. 3.**
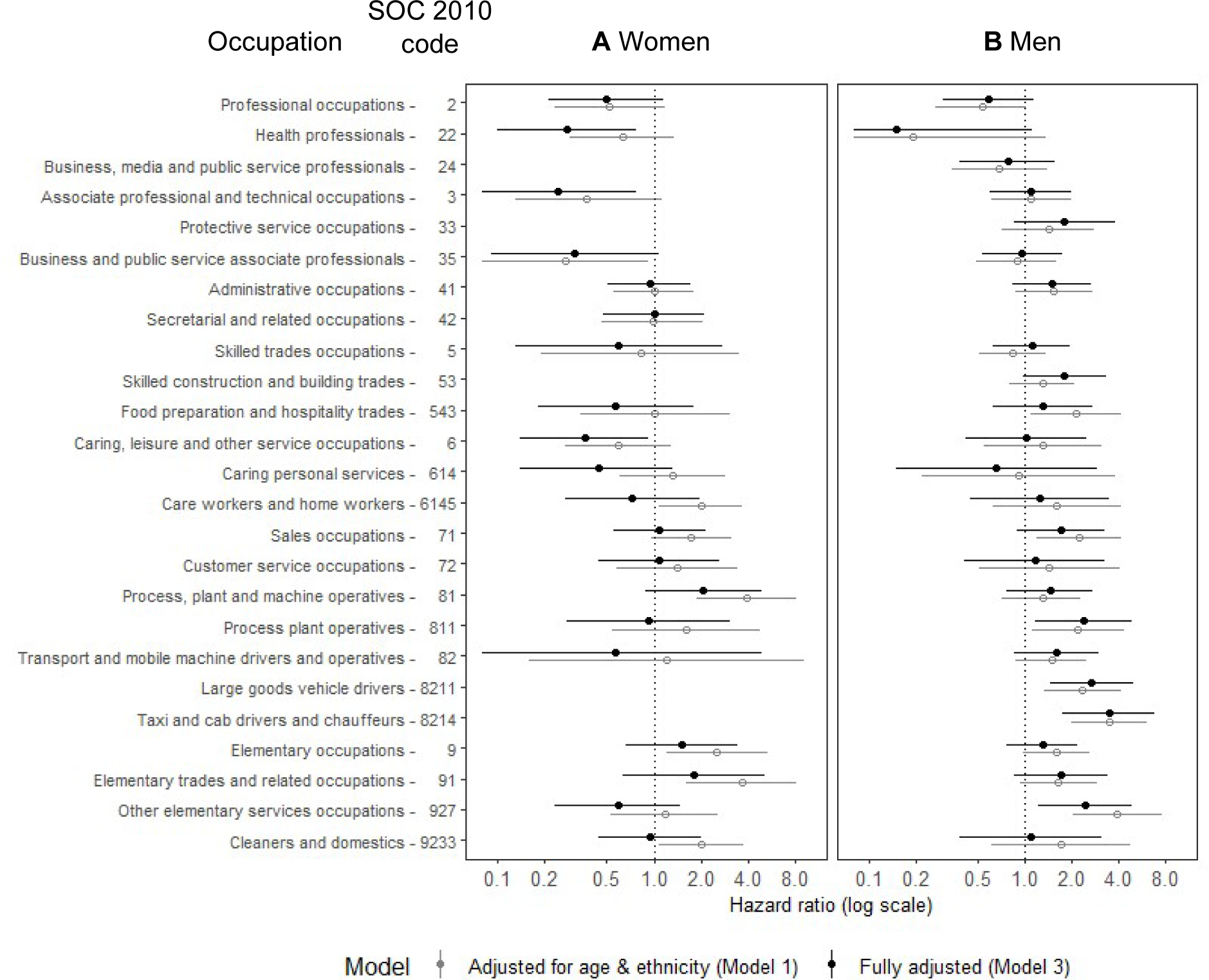
Hazard ratios for COVID-19 death for (A) women and (B) men aged 40-64 years in Scotland. _a_ Hazard ratios and 95% confidence intervals are displayed on a logarithmic scale; reference category: 1 ‘Managers, directors and senior officials’; fully-adjusted model (Model 3) is coloured in black (solid circle) and appears above Model 1 (grey hollow circle) for each group; fully-adjusted model (Model 3, Supplementary Table 7) additionally controls for: housing tenure, household size, household with children, whether multigenerational household, occupational exposure measures disease exposure, learning disability or difficulty, whether shielding and pre-pandemic health conditions; deaths occurring between 1 March 2020 and 31 January 2021; data are not reported if there were less than 10 deaths.

## Discussion

### Main results

Drawing on an innovative data collection, using Census, electronic health and residential data, we estimated COVID-19 hospital admission and death risks for different occupational groups, separately for women and men aged 40-64 years in Scotland between March 2020 to January 2021, i.e. before vaccination rollout. Generally, among women we found lower age-standardised COVID-19 admission and mortality rates. For both outcomes, the distribution of age-standardised rates largely presented an occupational gradient for both women and men, with lower rates among higher occupational groups and higher rates among lower occupational groups. Women working as process, plant and machine operatives (assemblers and sorters), and in elementary trades and related occupations (packers and canners) had the highest age-standardised mortality rates. Women working in caring personal services (nursing assistants/ambulance staff) also had high admission rates. Conversely, men in elementary services occupations (kitchen assistants/waiters), and taxi and cab drivers had the highest age-standardised mortality rates, with the latter group also having high admission rates. Previous studies covering a wider population in England and Wales reported similar results for COVID-19 mortality (11, 35).

Adjusting for socio-economic and pre-pandemic health factors reduced the association between the risk of COVID-19 hospital admission and death for most occupational groups, compared to the baseline model adjusting for basic demographic factors. Generally, we observed a similar occupational gradient for COVID-19 admission risks for women and men. Conversely, for COVID-19 death risks, the occupational gradients were different, with lower death risks among women in higher occupational groups (health professionals, associate professional and technical occupations, including paramedics and medical technicians), and higher death risks among men in lower occupational groups (taxi and large vehicle drivers, other elementary services occupations, including kitchen assistants and waiters, and food, drink and tobacco process plant operatives). These occupational groups may involve higher contact density, poorly ventilated workplaces, inability to work from home and higher financial strain which, when combined, greatly heighten risks.

### Strengths

To our knowledge, this is the first Scottish study to estimate COVID-19 hospital admission and death risks for detailed occupational groups among women and men, adjusting for a range of potential confounders. Indeed, it is one of only a few in the UK or elsewhere to combine such a range of socio-economic, health and household factors. First, we used a novel population-based data collection combining multiple data sources and covering a population of 1.7 million, which had sufficient power to identify COVID-19 hospitalisation and mortality risks associated with specific occupational groups among working-age women and men. Our results are likely to be relevant for other parts of the UK and other countries. Previous studies have examined these associations for the national population or in smaller populations [12, 14, 36]; other studies have reported only COVID-19 mortality risks [11, 35].

Second, we leveraged residential identifiers based on Unique Property Reference Numbers to construct a range of covariates, including household size, whether the household included children, and whether the household included multiple age-based generations. Household-level measures based on residential identifiers were used as potential confounders instead of measures based on 2011 Census data. This is considered an improvement compared to existing studies relying on Census-based measures [11, 19].

Third, in our analysis we adjusted for pre-pandemic health measures which were derived by combining long-term indicators from 2011 Census (e.g., whether the person had a learning disability or difficulty) and pre-pandemic health conditions, including cardiovascular conditions, diabetes, respiratory conditions and cancer/immunosuppression, derived using Read codes from primary care records collected from Scottish general practices. It is important to adjust for pre-pandemic health factors which can act as potential confounder. This was neglected in some studies [13, 36-38].

### Limitations

An important limitation of this study is that there may be some discrepancies which come from using occupational information from 2011 Census, as people may not be employed in the same occupation in March 2020 and subsequent changes during the pandemic. As a mitigation, we restricted the study population to those aged 40-64 years where occupational mobility is likely to be lower [11]. To corroborate this, we used data from the UK Household Longitudinal Study to show that the proportion of men and women who were in the same occupation between 2011 and 2020 was relatively high, suggesting that the Census occupation information was sufficiently stable over time. Our study estimates are largely consistent with both official estimates and other estimates from population-based studies using linked administrative data [11, 13, 35-37]. The occupational classification approach used in our study differs from previous investigations [13, 37], as we think it is important to distinguish between different groups of occupation within broader essential work clusters. Women and men may differ in the position and roles within the same occupational cluster.

Another limitation derives from the data linkage design. Our initial study population covers approximately 90% of the Scottish population. The linkage for this study relies on a population spine covering people who have interacted with the health services and received a Community Health Index (CHI) number. The population spine may not cover vulnerable population groups (refugees and migrants) that may be at an increased risk of severe COVID-19. Additional sources of bias derive from the linkage being conditional on having a valid 2011 Census record or a matched key for a UPRN residential identifier [23]. Misclassification may also arise due to errors in linked data and the data-linkage process [39]. We are aware that these sources of bias may affect our estimates but have insufficient information to ascertain how these differ from population-based estimates. Other limitations include the inability to investigate the role of changing restriction policies across the pandemic waves due to a limited study population. Determining the portion of the effect of occupation on COVID-19 outcomes attributable to occupational exposure and pre-pandemic health factors may require a mediation analysis approach which we leave for future research.

### Policy implications

Occupational differences in COVID-19 hospital admission and mortality risks between women and men may be explained by a combination of social, workplace and behavioural factors. These need to be considered when developing interventions to reduce gender discrepancies in any future COVID-19 waves or other respiratory epidemics. Enhancing the provision of appropriate protective equipment and training is required if we want to decrease the gap in adverse health outcomes between health professionals and those working in caring personal services (care and home workers). Coordinating social, workplace and behavioural interventions needs to be considered to reduce the transmission risks among more disadvantaged occupational groups, including taxi and large vehicle drivers, hospital porters, kitchen assistants and waiters, and workers in the food processing industry.

## Funding

This work was supported by UK Research Innovation Economic and Social Research Council (Grant numbers ES/W010321/1, ES/S007407/1 and ES/R005729/1). This research used data assets made available by National Safe Haven as part of the Data and Connectivity National Core Study, led by Health Data Research UK in partnership with the Office for National Statistics and funded by UK Research Innovation (Grant numbers MC_PC_20029 and MC_PC_20058).

## Ethics approval and data sharing

Ethical approval was obtained from the College of Social Sciences Research Ethics Committee of University of Glasgow (400200099). Approval for data linkage and its use for research was granted by the Scottish Public Benefit and Privacy Panel for Health and Social Care (2021-0119) and the Statistics Benefit and Privacy Panel (2021-0119). A protocol was submitted to both Public Benefit and Privacy Panels and is available upon request. The data used in this study are highly sensitive and will not be made publicly available.

## Data Availability

The data used in this study are highly sensitive and will not be made publicly available.

## Acknowledgments

A prior version of this paper was presented at the International Population Data Linkage (IPDLN) Conference 2022 in Edinburgh, UK, 7-9 September 2022 and published in its proceedings (40). We would like to acknowledge the contribution of Evan Williams (1990-2021) during an earlier stage of the research. We are also grateful to Gina Anghelescu for providing the code for the analysis conducted for Supplementary Table 2. We would like to thank colleagues from ADR-Scotland Data Acquisition Team, National Records of Scotland, and Public Health Scotland’s electronic Data Research and Innovation Services for their help and support during the development of this study.

## Supplementary material

**Table 1.** Occupational groups and SOC 2010 codes used in the study, including lower-level components (unit group, four-digit SOC 2010 codes)

| Occupational group | SOC 2010 codes | Lower-level components (unit group, four-digit SOC 2010 codes) |
| --- | --- | --- |
| Managers, directors and senior officials | 1 | 1115, 1116, 1121-1123, 1131-1136, 1139, 1150, 1161, 1162, 1171-1173, 1181, 1184, 1190, 1211, 1213, 1221, 1223-1226, 1241, 1242, 1251-1255, 1259 |
| Professional occupations | 2 | 2111-2114, 2119, 2121-2124, 2126, 2127, 2129, 2133-2137, 2139, 2141, 2142, 2150, 2311, 2312, 2314-2319 |
| Health professionals | 22 | 2211-2219, 2221-2223, 2229, 2231, 2232 |
| Business, media and public service professionals | 24 | 2412, 2413, 2419, 2421, 2423-2426, 2429, 2431-2436, 2442-2444, 2449, 2451, 2452, 2461-2463, 2471-2473 |
| Associate professional and technical occupations | 3 | 3111-3116, 3119, 3121, 3122, 3131, 3132, 3213, 3216-3219, 3231, 3233-3235, 3239, 3411-3417, 3421, 3422, 3441-3443 |
| Protective service occupations | 33 | 3311-3315, 3319 |
| Business and public service associate professionals | 35 | 3511-3513, 3520, 3531-3539, 3541-3546, 3550, 3561-3565, 3567 |
| Administrative occupations | 41 | 4112-4114, 4121-4124, 4129, 4131-4135, 4138, 4151, 4161, 4162 |
| Secretarial and related occupations | 42 | 4211-4217 |
| Skilled trades occupations | 5 | 5111-5114, 5119, 5211-5216, 5221-5225, 5231, 5232, 5234-5237, 5241, 5242, 5244, 5245, 5249, 5250, 5411-5414, 5419, 5421-5423, 5441-5443, 5449 |
| Skilled construction and building trades | 53 | 5311-5316, 5319, 5321-5323, 5330, |
| Food preparation and hospitality trades | 543 | 5431-5436 |
| Caring, leisure and other service occupations | 6 | 6121-6123, 6125, 6126, 6131, 6132, 6139, 6211, 6212, 6214, 6215, 6219, 6221, 6222, 6231, 6232, 6240 |
| Caring personal services | 614 | 6141-6144, 6146-6148 |
| Care workers and home workers | 6145 | 6145 |
| Sales occupations | 71 | 7111-7115, 7121-7125, 7129, 7130 |
| Customer service occupations | 72 | 7211, 7213-7215, 7219, 7220 |
| Process, plant and machine operatives | 81 | 8121-8127, 8129, 8131-8235, 8137, 8139, 8141-8143, 8149 |
| Process plant operatives | 811 | 8111-8119 |
| Transport and mobile machine drivers and operatives | 82 | 8212, 8213, 8215, 8221-8223, 8229, 8231-8234, 8239 |
| Large goods vehicle drivers | 8211 | 8211 |
| Taxi and cab drivers and chauffeurs | 8214 | 8214 |
| Elementary occupations | 9 | 9211, 9219, 9231-9236, 9239, 9241, 9242, 9249, 9251, 9259, 9260 |
| Elementary trades and related occupations | 91 | 9111, 9112, 9119, 9120, 9132, 9134, 9139 |
| Other elementary services occupations | 927 | 9271-9275, 9279 |
| Cleaners and domestics | 9244 | 9244 |

**Table 2.** Proportion of adults aged 40-64 years remaining in the same occupation between 2011-13 and 2018-19 in the UK, by major group occupation and sex.

| Major group occupation<br>(one-digit SOC 2010 code) | Population <sup>a</sup> |  |  | Women <sup>a</sup> |  |  | Men <sup>a</sup> |  |  |
| --- | --- | --- | --- | --- | --- | --- | --- | --- | --- |
|  | 2011-13<br>n | 2018-19<br>n | row<br>% <sup>b</sup> | 2011-13<br>n | 2018-19<br>n | row<br>% <sup>b</sup> | 2011-13<br>n | 2018-19<br>n | row<br>% <sup>b</sup> |
| 1 - Managers, directors and senior officials | 169 | 93 | 54.98 | 62 | 30 | 40.79 | 107 | 63 | 62.62 |
| 2 - Professional occupations | 290 | 202 | 70.91 | 152 | 111 | 73.18 | 138 | 91 | 68.76 |
| 3 - Associate professional and technical occupations | 261 | 142 | 54.23 | 101 | 52 | 53.10 | 160 | 90 | 54.82 |
| 4 - Administrative and secretarial occupations | 133 | 69 | 55.53 | 105 | 55 | 54.32 | 28 | 14 | 59.32 |
| 5 - Skilled trades occupations | 136 | 88 | 61.97 | 12 | 7 | 50.53 | 124 | 81 | 63.18 |
| 6 - Caring, leisure and other service occupations | 138 | 91 | 70.76 | 119 | 83 | 74.09 | 19 | 8 | 46.33 |
| 7 - Sales and customer service occupations | 104 | 55 | 50.83 | 73 | 42 | 56.59 | 31 | 13 | 38.79 |
| 8 - Process, plant and machine operatives | 109 | 72 | 68.30 | 14 | 6 | 46.19 | 95 | 66 | 71.97 |
| 9 - Elementary occupations | 127 | 71 | 60.48 | 68 | 38 | 65.22 | 59 | 33 | 54.87 |
| Overall | 1467 | 883 |  | 706 | 424 |  | 761 | 459 |  |

<sup>a</sup> The population includes all those who were present and employed at both time periods, and who were aged 40-64 years at wave 10, excluding those interviewed in 2020.
<sup>b</sup> Weighted percentages are reported.
Source: UKLHS wave 3 (2011-2013) and wave 10 (2018-2020).

**Table 3.** Covariates included in the statistical analysis.

| Variable | Coding/category description | Data source <sup>a</sup> |
| --- | --- | --- |
| *Socio-demographic variables:* |  |  |
| Age (years) | Restricted cubic spline with three knots (age 42, 53 and 62 years) | CHI register |
| Sex | Women, Men | CHI register |
| Ethnicity | Non-White, White | NRS 2011 Census |
| *Household-level variables:* |  |  |
| Housing tenure | Owned outright, Owned with mortgage, Social rented, Private rented, Owned/not known | NRS 2011 Census |
| Household size | 1-2 people, 3-4 people, 5-6 people, 7+ people | OS UPRN |
| Household with children | No children, At least one child 0-11 years, At least one child 12-17 years | CHI register and OS UPRN |
| Whether multigenerational household | Binary (Yes, No) variable identifying household with at least one person aged 65 or more years and a person at least 20 years younger | CHI register and OS UPRN |
| *Occupational exposure measures:* |  |  |
| Exposure to disease | Standardised score ranging from 0 (lowest frequency of exposure to disease or infection) to 1 (maximum frequency of exposure to disease or infection) | US DOL ETA O*NET survey data |
| Proximity to others | Standardised score ranging from 0 (no physical proximity to others) to 1 (maximum physical proximity to others) | US DOL ETA O*NET survey data |
| Environmentally controlled indoor conditions | Standardised score ranging from 0 (lowest frequency of working in environmentally controlled indoor conditions) to 1 (maximum frequency of working in environmentally controlled indoor conditions) | US DOL ETA O*NET survey data |
| *Health-related conditions:* |  |  |
| Learning disability or difficulty | Categorical variable (Yes, No, Not known) capturing whether a person has learning disability (e.g. Down's syndrome) or learning difficulty (e.g. dyslexia) | NRS 2011 Census |
| Whether shielding | Yes, No | PHS Shielding Patient List |
| Pre-pandemic health conditions | Binary (Yes, No) variables identifying whether a person has: cancer and immunosuppression, cardiovascular conditions, diabetes, hypertension, respiratory conditions, and other conditions | GP cluster data from PHS COVID-19 Research Database |

<sup>a</sup> Abbreviations: CHI: Community Health Index; NRS: National Records of Scotland; OS UPRN: Ordnance Survey Unique Property Reference Number; US DOL ETA: US Department of Labor, Employment and Training Administration; O\*NET: Occupational Information Network; PHS: Public Health Scotland; GP: General Practitioner.

**Table 4.** Description of the groups of people and conditions included in the shielding patient list.

| No. | Group |
| --- | --- |
| 1. | Recipients of solid organ transplant |
| 2. | People receiving chemotherapy or antibody treatment for cancer, including immunotherapy |
| 3. | People receiving radical radiotherapy for lung cancer |
| 4. | People receiving cancer treatments affecting immune system (protein kinase inhibitors or PARP inhibitors) |
| 5. | People with blood or bone marrow cancer (leukemia, lymphoma or myeloma) |
| 6. | Recipients of bone marrow or stem cell transplant in past 6 months or taking immunosuppressant medicine |
| 7. | People with severe lung condition (cystic fibrosis, severe asthma or COPD) |
| 8. | People with high risk of getting infections (SCID or sickle cell) |
| 9. | People taking medicine that makes them at higher risk of infections (steroids or immunosuppressant medicine) |
| 10. | People with serious heart condition or pregnant |
| 11. | People with problem with their spleen or spleen has been removed (splenectomy) |
| 12. | People with Down's syndrome |
| 13. | Recipient of dialysis or severe long-term kidney disease (stage 5) |

Source: Public Health Scotland. Search criteria for highest risk patients for inclusion to the shielding list. 2020. https://hpspubsrep.blob.core.windows.net/hps-website/nss/3008/documents/1_covid-19-search-criteria-highest-risk-patients.pdf. Accessed 15 Aug 2023.

**Table 5.**
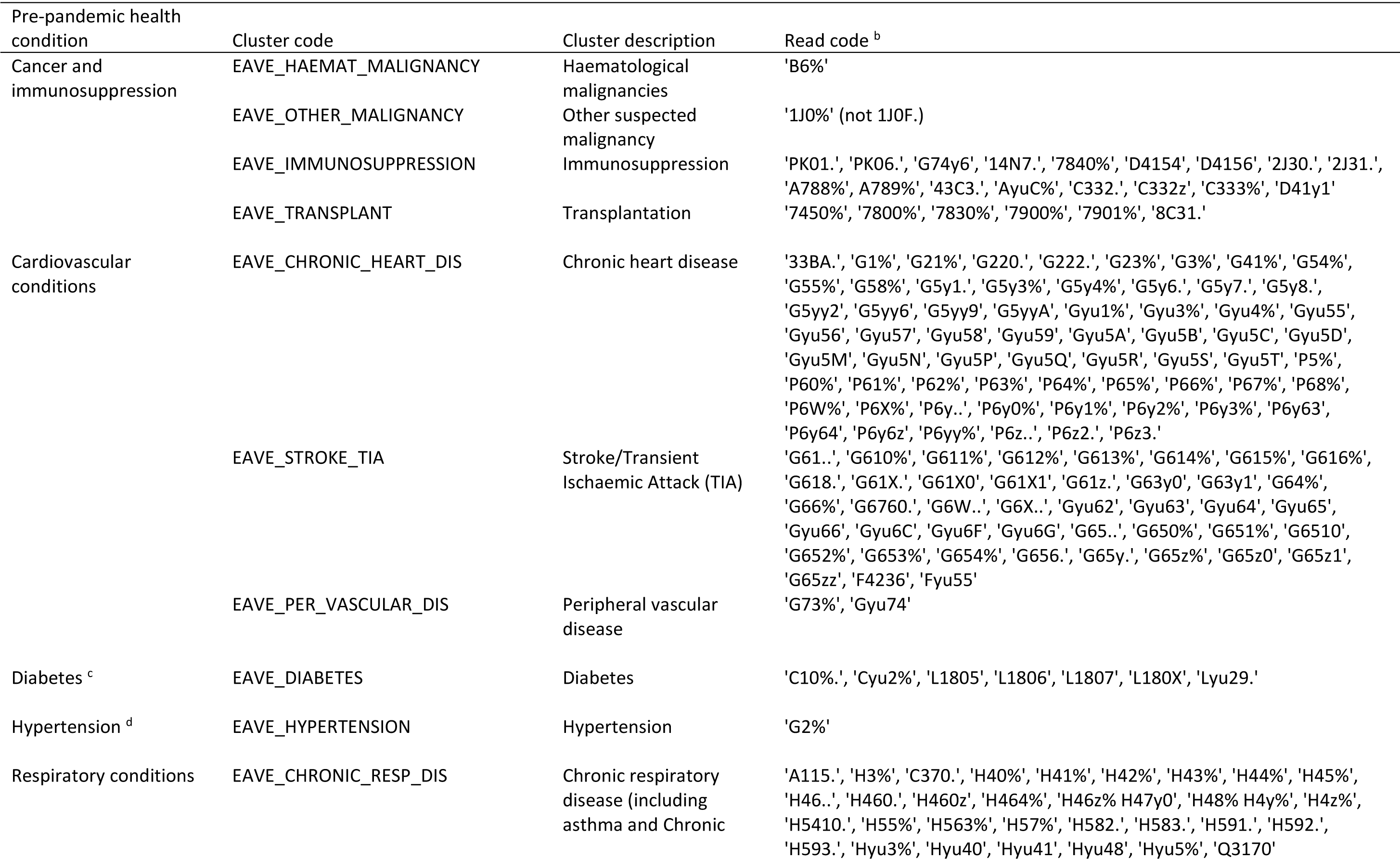
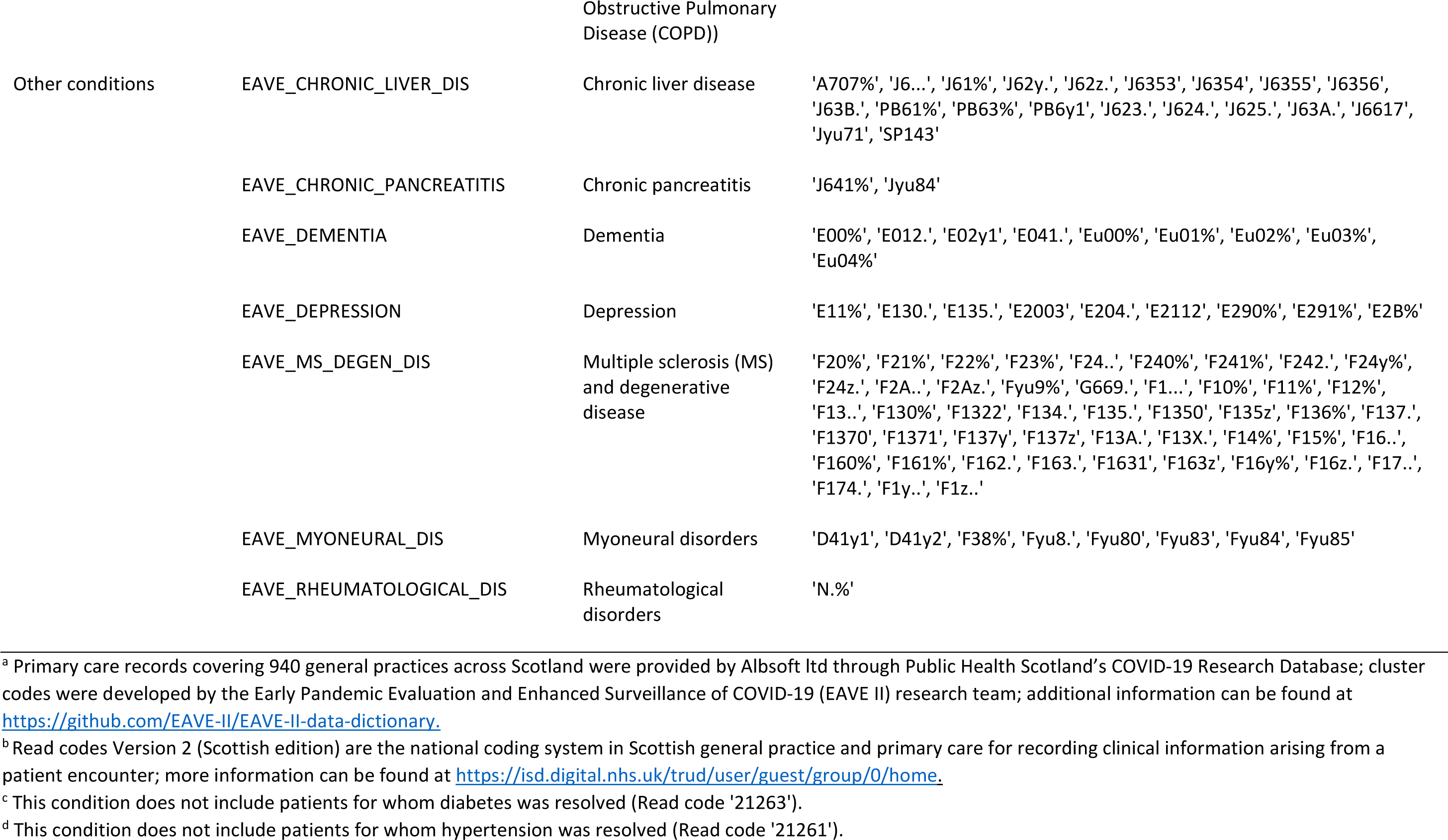
Clusters and Read codes used to derive pre-pandemic health conditions from primary care records ^a^.

**Table 6.** Full Cox proportional hazards models of risk of COVID-19 hospital admission for women and men aged 40-64 years in Scotland ^a^.

| Covariate | Model 1 |  | Model 2 |  | Model 3 |  |
| --- | --- | --- | --- | --- | --- | --- |
|  | Women | Men | Women | Men | Women | Men |
|  | HR (95%CI) <sup>b</sup> | HR (95%CI) <sup>b</sup> | HR (95%CI) <sup>b</sup> | HR (95%CI) <sup>b</sup> | HR (95%CI) <sup>b</sup> | HR (95%CI) <sup>b</sup> |
| *Age (years; restricted cubic spline):* |  |  |  |  |  |  |
| Age parameter 1 | **1.07 (1.05-1.09)** | **1.09 (1.07-1.10)** | **1.07 (1.05-1.09)** | **1.08 (1.07-1.10)** | **1.06 (1.04-1.07)** | **1.07 (1.05-1.09)** |
| Age parameter 2 | **0.97 (0.95-0.99)** | **0.98 (0.96-0.99)** | **0.97 (0.96-0.99)** | **0.98 (0.96-1.00)** | **0.97 (0.95-0.98)** | **0.97 (0.96-0.99)** |
| *Ethnicity (reference category: White):* |  |  |  |  |  |  |
| Not white | 1.09 (0.90-1.30) | **1.28 (1.06-1.52)** | **1.72 (1.43-2.06)** | **1.92 (1.63-2.27)** | **1.88 (1.57-2.25)** | **2.15 (1.84-2.52)** |
| *Occupation (SOC 2010 codes; reference category: 1 – Managers, directors and senior officials):* |  |  |  |  |  |  |
| 2 - Professional occupations | **0.58 (0.44-0.76)** | **0.49 (0.39-0.61)** | **0.52 (0.39-0.68)** | **0.49 (0.39-0.61)** | **0.55 (0.41-0.72)** | **0.52 (0.42-0.65)** |
| 22 - Health professionals | **1.63 (1.33-2.01)** | 1.25 (0.95-1.63) | 0.91 (0.67-1.22) | 0.85 (0.62-1.15) | 0.92 (0.68-1.24) | 0.87 (0.64-1.18) |
| 24 - Business, media and public service professionals | **0.49 (0.34-0.71)** | **0.61 (0.48-0.78)** | **0.50 (0.35-0.73)** | **0.63 (0.49-0.80)** | **0.52 (0.36-0.75)** | **0.66 (0.52-0.85)** |
| 3 - Associate professional and technical occupations | 0.83 (0.63-1.10) | 0.86 (0.69-1.06) | **0.63 (0.46-0.85)** | 0.81 (0.66-1.01) | **0.63 (0.46-0.85)** | 0.84 (0.67-1.04) |
| 33 - Protective service occupations | - | 1.24 (0.99-1.56) | - | 1.07 (0.82-1.39) | - | 1.12 (0.87-1.45) |
| 35 - Business and public service associate professionals | **0.60 (0.45-0.81)** | **0.66 (0.53-0.82)** | **0.63 (0.47-0.85)** | **0.67 (0.54-0.84)** | **0.65 (0.48-0.87)** | **0.69 (0.55-0.85)** |
| 41 - Administrative occupations | 1.06 (0.87-1.30) | 0.97 (0.78-1.22) | 1.00 (0.81-1.23) | 0.95 (0.76-1.19) | 1.00 (0.81-1.23) | 0.94 (0.75-1.18) |
| 42 - Secretarial and related occupations | 1.13 (0.88-1.45) | - | 1.10 (0.86-1.42) | - | 1.12 (0.88-1.44) | - |
| 5 - Skilled trades occupations | 0.85 (0.52-1.41) | 0.87 (0.74-1.03) | 0.79 (0.47-1.34) | 0.89 (0.73-1.09) | 0.75 (0.45-1.28) | 0.92 (0.75-1.12) |
| 53 - Skilled construction and building trades | - | 0.99 (0.84-1.18) | - | 0.96 (0.76-1.21) | - | 1.01 (0.80-1.27) |
| 543 - Food preparation and hospitality trades | **1.41 (1.01-1.96)** | 1.17 (0.90-1.52) | 1.07 (0.75-1.54) | 0.95 (0.72-1.27) | 0.99 (0.69-1.42) | 0.88 (0.66-1.17) |
| 6 - Caring, leisure and other service occupations | **1.25 (1.01-1.56)** | 1.21 (0.90-1.64) | 0.90 (0.68-1.18) | 1.04 (0.76-1.43) | 0.90 (0.68-1.19) | 1.01 (0.74-1.38) |
| 614 - Caring personal services | **2.26 (1.79-2.87)** | **2.45 (1.80-3.34)** | 1.13 (0.80-1.58) | **1.67 (1.18-2.37)** | 1.07 (0.76-1.51) | **1.57 (1.11-2.22)** |
| 6145 - Care workers and home workers | **2.16 (1.75-2.66)** | **1.97 (1.48-2.62)** | 1.24 (0.89-1.72) | **1.49 (1.08-2.06)** | 1.12 (0.80-1.56) | **1.38 (1.00-1.91)** |
| 71- Sales occupations | **1.43 (1.16-1.76)** | 1.27 (0.99-1.62) | 1.13 (0.89-1.43) | 1.13 (0.88-1.46) | 1.06 (0.84-1.34) | 1.06 (0.83-1.37) |
| 72 - Customer service occupations | **1.93 (1.47-2.54)** | 1.26 (0.88-1.79) | **1.72 (1.31-2.27)** | 1.18 (0.83-1.68) | **1.61 (1.22-2.11)** | 1.09 (0.77-1.56) |
| 81 - Process, plant and machine operatives | **2.20 (1.61-3.00)** | 1.05 (0.85-1.29) | **1.75 (1.24-2.49)** | 1.02 (0.80-1.29) | **1.49 (1.05-2.11)** | 1.02 (0.80-1.29) |
| 811 - Process plant operatives | **1.72 (1.18-2.51)** | 1.13 (0.84-1.52) | 1.38 (0.91-2.09) | 1.09 (0.80-1.49) | 1.24 (0.82-1.88) | 1.11 (0.82-1.51) |
| 82 - Transport and mobile machine drivers and operatives | 1.43 (0.75-2.72) | **1.34 (1.12-1.60)** | 1.02 (0.51-2.03) | 1.21 (0.97-1.51) | 0.92 (0.46-1.83) | 1.16 (0.93-1.45) |
| 8211 - Large goods vehicle drivers | - | **1.29 (1.01-1.63)** | - | 1.25 (0.97-1.62) | - | 1.24 (0.96-1.61) |
| 8214 - Taxi and cab drivers and chauffeurs | - | **2.37 (1.91-2.95)** | - | **2.04 (1.57-2.65)** | - | **1.84 (1.42-2.40)** |
| 9 - Elementary occupations | 1.21 (0.87-1.69) | **1.20 (1.00-1.43)** | 0.95 (0.67-1.35) | 1.06 (0.87-1.28) | 0.88 (0.62-1.26) | 1.03 (0.85-1.25) |
| 91 - Elementary trades and related occupations | **1.62 (1.10-2.38)** | 1.03 (0.82-1.29) | 1.22 (0.78-1.91) | 0.96 (0.73-1.26) | 1.08 (0.69-1.69) | 0.94 (0.72-1.24) |
| 927 - Other elementary services occupations | **1.56 (1.22-2.00)** | **1.59 (1.18-2.15)** | 1.11 (0.83-1.49) | 1.32 (0.96-1.81) | 1.02 (0.76-1.37) | 1.20 (0.88-1.64) |
| 9233 - Cleaners and domestics | **1.90 (1.52-2.37)** | 0.90 (0.57-1.44) | 1.31 (1.00-1.73) | 0.72 (0.45-1.15) | 1.16 (0.88-1.53) | 0.68 (0.42-1.08) |
| No code required | **1.82 (1.43-2.32)** | 0.94 (0.76-1.16) | **2.36 (1.30-4.29)** | **2.39 (1.67-3.42)** | 1.77 (0.98-3.22) | **2.20 (1.54-3.13)** |
| *Housing tenure (reference category: Owned outright):* |  |  |  |  |  |  |
| Owned with mortgage |  |  | **1.42 (1.24-1.61)** | **1.47 (1.30-1.66)** | **1.34 (1.18-1.53)** | **1.43 (1.27-1.62)** |
| Social rented |  |  | **2.15 (1.87-2.48)** | **1.99 (1.73-2.28)** | **1.58 (1.37-1.83)** | **1.57 (1.37-1.81)** |
| Private rented |  |  | **1.30 (1.08-1.58)** | **1.23 (1.03-1.47)** | 1.12 (0.93-1.36) | 1.11 (0.93-1.33) |
| Other/not known |  |  | **0.59 (0.44-0.78)** | **0.32 (0.24-0.43)** | **2.39 (1.41-4.02)** | **1.91 (1.33-2.74)** |
| *Household size (UPRN; reference category: 1-2 people):* |  |  |  |  |  |  |
| 3-4 people |  |  | 0.97 (0.89-1.06) | **1.11 (1.02-1.20)** | 1.01 (0.92-1.10) | **1.14 (1.05-1.24)** |
| 5-6 people |  |  | 1.06 (0.92-1.23) | **1.25 (1.09-1.43)** | 1.11 (0.96-1.29) | **1.27 (1.11-1.46)** |
| 7+ people |  |  | **1.54 (1.23-1.94)** | **2.07 (1.72-2.48)** | **1.51 (1.19-1.90)** | **1.92 (1.59-2.31)** |
| *Household with children (UPRN; reference category: No children):* |  |  |  |  |  |  |
| At least one child (0-11 years) |  |  | 0.91 (0.77-1.06) | **0.81 (0.70-0.94)** | 0.90 (0.77-1.05) | **0.83 (0.72-0.96)** |
| At least one child (12-17 years) |  |  | 0.93 (0.82-1.06) | **0.83 (0.73-0.95)** | 0.94 (0.82-1.07) | **0.85 (0.75-0.97)** |
| *Whether multigenerational household (UPRN; reference category: No):* |  |  |  |  |  |  |
| Yes |  |  | 1.03 (0.89-1.18) | 1.06 (0.93-1.21) | 0.99 (0.86-1.14) | 1.02 (0.90-1.17) |
| *Occupational exposure measures (O\*NET scores 0-1):* |  |  |  |  |  |  |
| Exposure to disease |  |  | **1.83 (1.44-2.33)** | **1.61 (1.38-1.88)** | **1.81 (1.42-2.31)** | **1.67 (1.43-1.95)** |
| Proximity to others |  |  | **1.75 (1.05-2.94)** | 1.17 (0.81-1.67) | **1.71 (1.03-2.87)** | 1.06 (0.74-1.52) |
| Environmentally controlled indoor conditions |  |  | 0.83 (0.45-1.53) | 1.09 (0.83-1.45) | 0.81 (0.44-1.50) | 1.09 (0.82-1.44) |
| *Learning disability or difficulty (reference category: No):* |  |  |  |  |  |  |
| Yes |  |  |  |  | **1.44 (1.15-1.81)** | **1.42 (1.17-1.72)** |
| Not known |  |  |  |  | **0.25 (0.14-0.43)** | **0.14 (0.09-0.21)** |
| *Whether shielding (reference category: No):* |  |  |  |  |  |  |
| Yes |  |  |  |  | **3.24 (2.86-3.68)** | **3.26 (2.83-3.76)** |
| *Pre-pandemic health conditions (reference category for each covariate: No):* |  |  |  |  |  |  |
| Cancer and immunosuppression Yes |  |  |  |  | **1.66 (1.36-2.04)** | **1.46 (1.18-1.80)** |
| Cardiovascular conditions |  |  |  |  | **1.53 (1.36-1.71)** | **1.58 (1.43-1.73)** |
| Diabetes |  |  |  |  | **2.25 (2.02-2.51)** | **2.16 (1.97-2.37)** |
| Hypertension |  |  |  |  | **1.44 (1.31-1.58)** | **1.37 (1.26-1.49)** |
| Respiratory conditions |  |  |  |  | **1.40 (1.28-1.54)** | **1.16 (1.05-1.28)** |
| Other conditions |  |  |  |  | **1.36 (1.25-1.47)** | **1.38 (1.26-1.50)** |
| COVID-19 hospital admissions (n) | 2,820 | 3,140 | 2,820 | 3,140 | 2,820 | 3,140 |
| Total persons (n) | 859,800 | 840,870 | 859,800 | 840,870 | 859,800 | 840,870 |
| Total episodes (n) | 937,810 | 914,600 | 937,810 | 914,600 | 937,810 | 914,600 |
| AIC | 76,175 | 84,755 | 79,955 | 84,435 | 74,871 | 83,392 |
| BIC | 76,504 | 85,095 | 76,437 | 84,928 | 75,471 | 83,990 |

<sup>a</sup> Hospital admissions occurring between 1 March 2020 and 31 January 2021.
<sup>b</sup> Hazard Ratios (HR) and 95% Confidence Intervals (CI) are reported; estimates are not reported if there were less than 10 hospital admissions; HRs above 1 with $p < 0.05$ are highlighted in bold and HRs below 1 with $p < 0.05$ are highlighted in white (grey cells).

**Table 7.** Full Cox proportional hazards models of risk of COVID-19 related death for women and men aged 40-64 years in Scotland ^a^.

| Covariate | Model 1 |  | Model 2 |  | Model 3 |  |
| --- | --- | --- | --- | --- | --- | --- |
|  | Women | Men | Women | Men | Women | Men |
|  | HR (95%CI) <sup>b</sup> | HR (95%CI) <sup>b</sup> | HR (95%CI) <sup>b</sup> | HR (95%CI) <sup>b</sup> | HR (95%CI) <sup>b</sup> | HR (95%CI) <sup>b</sup> |
| *Age (years; restricted cubic spline):* |  |  |  |  |  |  |
| Age parameter 1 | **1.08 (1.02-1.14)** | **1.13 (1.07-1.19)** | **1.08 (1.02-1.15)** | **1.13 (1.07-1.19)** | 1.06 (0.99-1.12) | **1.11 (1.05-1.17)** |
| Age parameter 2 | 1.02 (0.96-1.08) | 1.01 (0.96-1.06) | 1.01 (0.96-1.07) | 1.01 (0.96-1.06) | 1.01 (0.95-1.06) | 1.00 (0.95-1.05) |
| *Ethnicity (reference category: White):* |  |  |  |  |  |  |
| Not white | **0.43 (0.24-0.75)** | **0.34 (0.22-0.52)** | 1.25 (0.68-2.29) | 0.72 (0.41-1.25) | 1.57 (0.88-2.80) | **1.87 (1.18-2.96)** |
| *Occupation (SOC 2010 codes; reference category: 1 – Managers, directors and senior officials):* |  |  |  |  |  |  |
| 2 - Professional occupations | 0.52 (0.23-1.17) | 0.53 (0.27-1.01) | 0.44 (0.19-1.04) | 0.54 (0.28-1.03) | 0.49 (0.21-1.15) | 0.59 (0.30-1.13) |
| 22 - Health professionals | 0.63 (0.29-1.35) | 0.19 (0.03-1.37) | **0.27 (0.10-0.75)** | 0.14 (0.02-1.08) | **0.28 (0.10-0.78)** | 0.15 (0.02-1.11) |
| 24 - Business, media and public service professionals | - | 0.68 (0.34-1.38) | - | 0.71 (0.35-1.43) | - | 0.78 (0.38-1.57) |
| 3 - Associate professional and technical occupations | 0.37 (0.13-1.12) | 1.00 (0.61-1.98) | **0.24 (0.07-0.77)** | 1.03 (0.57-1.89) | **0.24 (0.07-0.78)** | 1.10 (0.60-2.01) |
| 33 - Protective service occupations | - | 1.42 (0.72-2.79) | - | 1.63 (0.77-3.46) | - | 1.79 (0.85-3.78) |
| 35 - Business and public service associate professionals | **0.27 (0.08-0.92)** | 0.89 (0.49-1.61) | 0.30 (0.09-1.02) | 0.91 (0.51-1.65) | 0.31 (0.09-1.07) | 0.96 (0.53-1.74) |
| 41 - Administrative occupations | 1.00 (0.55-1.82) | 1.54 (0.87-2.72) | 0.93 (0.50-1.71) | 1.45 (0.82-2.58) | 0.94 (0.51-1.73) | 1.48 (0.83-2.64) |
| 42 - Secretarial and related occupations | 0.98 (0.46-2.07) | - | 0.95 (0.45-2.02) | - | 1.00 (0.47-2.12) | - |
| 5 - Skilled trades occupations | 0.82 (0.19-3.55) | 0.83 (0.51-1.35) | 0.67 (0.14-3.13) | 1.05 (0.60-1.86) | 0.59 (0.13-2.75) | 1.12 (0.63-1.97) |
| 53 - Skilled construction and building trades | - | 1.29 (0.80-2.08) | - | 1.66 (0.89-3.08) | - | 1.80 (0.97-3.34) |
| 543 - Food preparation and hospitality trades | 1.02 (0.34-3.07) | **2.12 (1.08-4.18)** | 0.64 (0.20-2.08) | 1.56 (0.75-3.26) | 0.57 (0.18-1.82) | 1.31 (0.63-2.73) |
| 6 - Caring, leisure and other service occupations | 0.59 (0.27-1.29) | 1.30 (0.55-3.08) | **0.35 (0.13-0.91)** | 1.07 (0.44-2.58) | **0.36 (0.14-0.92)** | 1.02 (0.42-2.47) |
| 614 - Caring personal services | 1.32 (0.60-2.90) | 0.92 (0.22-3.83) | 0.46 (0.15-1.39) | 0.70 (0.16-3.10) | 0.44 (0.14-1.32) | 0.66 (0.15-2.92) |
| 6145 - Care workers and home workers | **2.00 (1.08-3.69)** | 1.61 (0.63-4.12) | 0.83 (0.30-2.27) | 1.36 (0.49-3.80) | 0.73 (0.27-1.98) | 1.26 (0.45-3.51) |
| 71- Sales occupations | 1.73 (0.96-3.13) | **2.21 (1.19-4.12)** | 1.19 (0.60-2.37) | 1.86 (0.97-3.56) | 1.09 (0.55-2.15) | 1.71 (0.89-3.26) |
| 72 - Customer service occupations | 1.42 (0.58-3.44) | 1.44 (0.51-4.04) | 1.19 (0.49-2.92) | 1.24 (0.44-3.50) | 1.08 (0.44-2.64) | 1.16 (0.41-3.26) |
| 81 - Process, plant and machine operatives | **3.96 (1.90-8.24)** | 1.29 (0.72-2.30) | **2.67 (1.13-6.33)** | 1.43 (0.76-2.70) | 2.08 (0.88-4.95) | 1.45 (0.77-2.74) |
| 811 - Process plant operatives | 1.60 (0.54-4.80) | **2.19 (1.11-4.30)** | 1.08 (0.32-3.59) | **2.32 (1.13-4.76)** | 0.92 (0.28-3.07) | **2.39 (1.16-4.91)** |
| 82 - Transport and mobile machine drivers and operatives | 1.22 (0.16-9.17) | 1.48 (0.88-2.47) | 0.66 (0.08-5.70) | 1.61 (0.86-3.02) | 0.57 (0.07-4.93) | 1.60 (0.86-3.00) |
| 8211 - Large goods vehicle drivers | - | **2.34 (1.32-4.13)** | - | **2.63 (1.42-4.88)** | - | **2.69 (1.45-4.99)** |
| 8214 - Taxi and cab drivers and chauffeurs | - | **3.48 (1.99-6.08)** | - | **3.77 (1.90-7.48)** | - | **3.46 (1.74-6.86)** |
| 9 - Elementary occupations | **2.53 (1.20-5.35)** | 1.59 (0.97-2.60) | 1.68 (0.74-3.82) | 1.31 (0.77-2.22) | 1.51 (0.66-3.43) | 1.30 (0.77-2.20) |
| 91 - Elementary trades and related occupations | **3.65 (1.61-8.26)** | 1.64 (0.93-2.90) | 2.16 (0.76-6.16) | 1.74 (0.87-3.50) | 1.80 (0.63-5.11) | 1.70 (0.85-3.42) |
| 927 - Other elementary services occupations | 1.18 (0.53-2.59) | **3.90 (2.03-7.52)** | 0.67 (0.26-1.70) | **2.80 (1.39-5.65)** | 0.59 (0.23-1.49) | **2.44 (1.21-4.91)** |
| 9233 - Cleaners and domestics | **2.02 (1.07-3.80)** | 1.70 (0.61-4.79) | 1.14 (0.53-2.43) | 1.22 (0.42-3.52) | 0.95 (0.44-2.03) | 1.09 (0.38-3.13) |
| No code required | **3.78 (1.96-7.27)** | **5.91 (3.61-9.67)** | 3.36 (0.68-16.56) | **7.45 (2.93-18.95)** | 2.02 (0.41-10.00) | **6.17 (2.47-15.41)** |
| *Housing tenure (reference category: Owned outright):* |  |  |  |  |  |  |
| Owned with mortgage |  |  | 1.19 (0.82-1.75) | **1.59 (1.14-2.22)** | 1.12 (0.76-1.63) | **1.55 (1.11-2.17)** |
| Social rented |  |  | **2.47 (1.67-3.66)** | **2.93 (2.06-4.18)** | **1.59 (1.07-2.37)** | **2.12 (1.48-3.03)** |
| Private rented |  |  | 1.51 (0.87-2.60) | **1.79 (1.12-2.84)** | 1.22 (0.71-2.11) | 1.50 (0.94-2.40) |
| Other/not known |  |  | **0.30 (0.13-0.71)** | 0.82 (0.41-1.67) | 1.27 (0.36-4.46) | **3.06 (1.62-5.77)** |
| *Household size (UPRN; reference category: 1-2 people):* |  |  |  |  |  |  |
| 3-4 people |  |  | **0.74 (0.55-0.98)** | 0.92 (0.74-1.16) | 0.78 (0.58-1.04) | 0.96 (0.77-1.21) |
| 5-6 people |  |  | 0.65 (0.36-1.17) | 0.95 (0.61-1.47) | 0.68 (0.38-1.23) | 0.94 (0.60-1.46) |
| 7+ people |  |  | **3.03 (1.77-5.19)** | **4.87 (3.37-7.05)** | **2.84 (1.63-4.96)** | **3.78 (2.54-5.61)** |
| *Household with children (UPRN; reference category: No children):* |  |  |  |  |  |  |
| At least one child (0-11 years) |  |  | 0.61 (0.33-1.12) | **0.35 (0.19-0.64)** | 0.60 (0.33-1.12) | **0.37 (0.21-0.68)** |
| At least one child (12-17 years) |  |  | 0.81 (0.49-1.34) | **0.64 (0.41-0.98)** | 0.82 (0.50-1.35) | 0.68 (0.44-1.04) |
| *Whether multigenerational household (UPRN; reference category: No):* |  |  |  |  |  |  |
| Yes |  |  | **1.91 (1.33-2.74)** | **1.77 (1.32-2.36)** | **1.80 (1.25-2.60)** | **1.65 (1.22-2.23)** |
| *Occupational exposure measures (O\*NET scores 0-1):* |  |  |  |  |  |  |
| Exposure to disease |  |  | **2.41 (1.22-4.73)** | 1.40 (0.94-2.10) | **2.38 (1.21-4.71)** | **1.51 (1.01-2.26)** |
| Proximity to others |  |  | 2.57 (0.47-14.20) | 1.00 (0.36-2.73) | 2.40 (0.44-13.10) | 0.80 (0.30-2.16) |
| Environmentally controlled indoor conditions |  |  | 0.70 (0.14-3.56) | **2.48 (1.15-5.32)** | 0.66 (0.13-3.42) | **2.47 (1.16-5.29)** |
| *Learning disability or difficulty (reference category: No):* |  |  |  |  |  |  |
| Yes |  |  |  |  | **2.01 (1.20-3.36)** | **1.58 (1.05-2.37)** |
| Not known |  |  |  |  | **0.21 (0.05-0.92)** | **0.09 (0.04-0.20)** |
| *Whether shielding (reference category: No):* |  |  |  |  |  |  |
| Yes |  |  |  |  | **4.95 (3.57-6.86)** | **4.95 (3.68-6.66)** |
| *Pre-pandemic health conditions (reference category for each covariate: No):* |  |  |  |  |  |  |
| Cancer and immunosuppression |  |  |  |  | 1.66 (0.96-2.87) | 1.49 (0.94-2.38) |
| Cardiovascular conditions |  |  |  |  | **1.89 (1.42-2.53)** | **1.64 (1.31-2.05)** |
| Diabetes |  |  |  |  | **2.98 (2.25-3.95)** | **2.13 (1.70-2.67)** |
| Hypertension |  |  |  |  | 1.19 (0.91-1.56) | 1.19 (0.96-1.47) |
| Respiratory conditions |  |  |  |  | 1.14 (0.85-1.52) | 1.04 (0.81-1.34) |
| Other conditions |  |  |  |  | **1.45 (1.14-1.84)** | **1.80 (1.46-2.21)** |
| COVID-19 deaths (n) | 300 | 460 | 300 | 460 | 300 | 460 |
| Total persons (n) | 859,800 | 840,870 | 859,800 | 840,870 | 859,800 | 840,870 |
| AIC | 7,968 | 12,189 | 7,885 | 12,031 | 7,658 | 11,744 |
| BIC | 8,260 | 12,527 | 8,328 | 12,520 | 8,206 | 12,338 |

<sup>a</sup> Deaths occurring between 1 March 2020 and 31 January 2021.
<sup>b</sup> Hazard Ratios (HR) and 95% Confidence Intervals (CI) are reported; estimates are not reported if there were less than 10 deaths; HRs above 1 with $p < 0.05$ are highlighted in bold and HRs below 1 with $p < 0.05$ are highlighted in white (grey cells).

## Notes

### Competing Interest Statement

The authors have declared no competing interest.

### Funding Statement

The study was funded by UK Research Innovation Economic and Social Research Council (Grant numbers ES/W010321/1, ES/S007407/1 and ES/R005729/1).
The study used data assets made available by National Safe Haven as part of the Data and Connectivity National Core Study, led by Health Data Research UK in partnership with the Office for National Statistics and funded by UK Research Innovation (Grant numbers MC_PC_20029 and MC_PC_20058).

### Author Declarations

The College of Social Sciences Research Ethics Committee of University of Glasgow gave ethical approval (400200099). The Scottish Public Benefit and Privacy Panel for Health and Social Care and the Statistics Benefit and Privacy Panel gave approval for data linkage (2021-0119).

## References

1. Peckham H, de Gruijter NM, Raine C, Radziszewka A, Ciurtin C, Wedderburn LR, et al. Male sex identified by global COVID-19 meta-analysis as a risk factor for death and ITU admission. Nat Commun. 2020; 10.1038/s41467-020-19741-6.

2. Doerre A, Doblhammer, G. The influence of gender on COVID-19 infections and mortality in Germany: Insights from age- and gender-specific modelling of contact rates, and deaths in the early phase of the pandemic. PloS One. 2022; 10.1371/journal.pone.0268119.

3. Sobotka T, Brzozowska Z, Muttarak R, Zeman K, Di Lego V. Age, gender and COVID-19 infections. MedRxiv. 2020. 10.1101/2020.05.24.201111765.

4. Scully EP, Schumock G, Fu M, Massaccesi G, Muschelli J, Betz J, et al. Sex and gender differences in testing, hospital admission, clinical presentation, and drivers of severe outcomes from COVID-19. Open Forum Infect Dis. 2021; 10.1093/ofid/ofab448.

5. Stalter RM, Atluri V, Xia F, Thomas KK, Lan KF, Greninger AL, et al. Elucidating pathways mediating the relationship between male sex and COVID-19 severity. Clin Epidemiol. 2022; 10.2147/CLEP.S335494.

6. Hâncean M-G, Lerner J, Perc M, Oană I, Bunaciu D-A, Stoica AA, et al. Occupations and their impact on the spreading of COVID-19 in urban communities. Sci Rep. 2022; 10.1038/s41598-022-18392-5.

7. Oude Hengel KM, Burdorf A, Pronk A, Schlünssen V, Stokholm ZA, Kolstad HA, et al. Exposure to SARS-CoV-2 infection at work: development of an international job exposure matrix (COVID-19-JEM). Scand J Work Environ Health. 2022; 10.5271/sjweh.3998.

8. Albanesi S, Kim J. Effects of the COVID-19 recession on the US labor market: occupation, family, and gender. JEP. 2021; 10.1257/jep.35.3.3.

9. Blau FD, Koebe J, Meyerhofer PA. Who are the essential and frontline workers? Bus Econ. 2021; 10.1057/s11369-021-00230-7.

10. Beale S, Patel P., Rodger A, Braithwite I, Byrne T, Fong WLE, et al. Occupation, work-related contact and SARS-CoV-2 anti-nucleocapsid serological status: findings from the Virus Watch prospective cohort study. Occup Environ Med. 2022; 10.1136/oemed-2021-107920.

11. Nafilyan V, Pawelek P, Ayoubkhani D, Rhodes S, Pembrey L, Matz M, et al. Occupation and COVID-19 mortality in England: a national linked data study of 14.3 million adults. Occup Environ Med. 2021; 10.1136/oemed-2021-107818.

12. Mutambudzi M, Niedzwiedz C, Macdonald EB, Leyland A, Mair F, Anderson J et al. Occupation and risk of severe COVID-19: prospective cohort study of 120 075 UK Biobank participants. Occup Environ Med. 2020; https://oem.bmj.com/content/78/5/307.

13. Cherrie M, Rhodes S, Wilkinson J, Mueller W, Nafilyan V, Van Tongeren M, et al. Longitudinal changes in proportionate mortality due to COVID-19 by occupation in England and Wales. Scand J Work Environ Health. 2022; https://www.sjweh.fi/article/4048.

14. Fenton L, Gribben C, Caldwell D, Colville S, Bishop J, Reid M, et al. Risk of hospital admission with covid-19 among teachers compared with healthcare workers and other adults of working age in Scotland, March 2020 to July 2021: population based case-control study. BMJ. 2021; 10.1136/bmj.n2060.

15. Galasso V, Pons V, Profeta P, Becher M, Brouard S, Foucault M. Gender differences in COVID-19 attitudes and behavior: panel evidence from eight countries. PNAS 2020; https://www.pnas.org/doi/full/10.1073/pnas.2012520117.

16. Abate BB, Kassie AM, Kassaw MW, Aragie TG, Masresha SA. Sex difference in coronavirus disease (COVID-19): a systematic review and meta-analysis. BMJ Open. 2020; 10.1136/bmjopen-2020-040129.

17. Bambra C, Riordan R, Ford J, Matthews F. The COVID-19 pandemic and health inequalities. J Epidemiol Community Health. 2020; 10.1136/jech-2020-214401.

18. Williamson EJ, Walker AJ, Bhaskaran K, Bacon S, Bates C, Morton CE, et al. Factors associated with COVID-19-related death using OpenSAFELY. Nature. 2020; 10.1038/s41586-020-2521-4.

19. Nafilyan V, Islam N, Ayoubkhani D, Gilles C, Katikireddi SV, Mathur R, et al. Ethnicity, household composition and COVID-19 mortality: a national linked data study. J R Soc Med. 2021; 10.1177/0141076821999973.

20. Public Health Scotland. COVID-19 Data for Research. 2020. https://www.isdscotland.org/Products-and-Services/EDRIS/COVID-19/. Accessed 15 Jun 2023.

21. Scottish Centre for Administrative Data Research. Investigating socioeconomic, household and environmental risk factors for Covid-19 in Scotland. 2021. https://www.scadr.ac.uk/our-research/health-and-social-care/investigating-socioeconomic-household-and-environmental-risk. Accessed 15 Jun 2023.

22. Scottish Government. Scotland’s Data and Intelligence Network. 2020. https://www.gov.scot/groups/data-and-intelligence-network/. Accessed 15 Jun 2023.

23. Clark D, Dibben C. A guide to CHI-UPRN Residential Linkage (CURL) file. Administrative Data Research Scotland (ADR Scotland). 2020. https://www.scadr.ac.uk/our-research/innovations/addressing-people-scotland-linking-chi-and-uprn. Accessed 15 Jun 2023.

24. National Records of Scotland. Mid-year population estimates. Scotland, mid-2019. 2020. https://www.nrscotland.gov.uk/files/statistics/population-estimates/mid-19/mid-year-pop-est-19-report.pdf. Accessed 15 Jun 2023.

25. Scottish Government. Coronavirus (COVID-19) data: definitions and sources. 2020. https://www.gov.scot/publications/coronavirus-covid-19-data-definitions-and-sources/#Published%20data%20on%20COVID-19%20cases,%20deaths%20and%20testing. Accessed 15 Jun 2023.

26. Office for National Statistics. SOC 2010. 2022. https://www.ons.gov.uk/methodology/classificationsandstandards/standardoccupationalclassificationsoc/soc2010. Accessed 15 Aug 2022.

27. University of Essex, Institute for Social and Economic Research. Understanding Society: Waves 1-11, 2009-2020 and Harmonised BHPS: Waves 1-18, 1991-2009. 2022 15th Edition. UK Data Service. 10.5255/ukda-sn-6614-16. Accessed 12 May 2022.

28. US Department of Labor, Employment and Training Administration. O*NET 24.2 Data Dictionary. 2020. https://www.onetcenter.org/dictionary/24.2/excel/. Accessed 7 May 2021.

29. Office for National Statistics. Which occupations have the highest potential exposure to the coronavirus (COVID-19)? 2020. https://www.ons.gov.uk/employmentandlabourmarket/peopleinwork/employmentandemployeetypes/articles/whichoccupationshavethehighestpotentialexposuretothecoronaviruscovid19/2020-05-11. Accessed 23 Jun 2021.

30. Clift AK, Coupland CA, Keogh RH, Diaz-Ordaz K, Williamson E, Harrison EM, et al. Living risk prediction algorithm (QCOVID) for risk of hospital admission and mortality from coronavirus 19 in adults: national derivation and validation cohort study. BMJ. 2020; 10.1136/bmj.m3731.

31. Eurostat. Revision of the European Standard Population-Report of Eurostat’s task force. Luxembourg: Publication office of the European Union; 2013; 10.2785/11470. Accessed 13 Jun 2021.

32. Dobson AJ, Kuulasmaa K, Eberle E, Scherer J. Confidence intervals for weighted sums of poisson parameters. Stat Med. 1991; 10.1002/sim.4780100317.

33. Consonni D, Coviello E, Buzzoni C, Mensi C. A command to calculate age-standardized rates with efficient interval estimation. SJ. 2012; 10.1177/1536867X1201200408.

34. Morris JK, Tan J, Fryers P, Bestwick J. Evaluation of stability of directly standardized rates for sparse data using simulation methods. PJM. 2018; 10.1186/s12963-018-0177-1.

35. Office for National Statistics. Coronavirus (COVID-19) related deaths by occupation, England and Wales: deaths registered between 9 March and 28 December 2020. 2021. https://www.ons.gov.uk/peoplepopulationandcommunity/healthandsocialcare/causesofdeath/bulletins/coronaviruscovid19relateddeathsbyoccupationenglandandwales/latest#women-and-deaths-involving-covid-19-by-occupation. Accessed 13 Jun 2023.

36. Magnusson K, Nygård K, Methi F, Vold L, Telle K. Occupational risk of COVID-19 in the first versus second epidemic wave in Norway, 2020. Euro Surveill. 2021; 10.2807/1560-7917.ES.2021.26.40.2001875.

37. Billingsley S, Brandén M, Aradhya S, Drefahl S, Andersson G, Mussino E. COVID-19 mortality across occupations and secondary risks for elderly individuals in the household: a population register-based study. Scand J Work Environ Health. 2022; 10.5271/sjweh.3992.

38. Drefahl S, Wallace M, Mussino E, Aradhya S, Kolk M, Brandén M, et al. A population-based cohort study of socio-demographic risk factors for COVID-19 deaths in Sweden. Nat Commun 2020; 10.1038/s41467-020-18926-3.

39. Shaw RJ, Harron KL, Pescarini JM, Pinto Junior EP, Allik M, Siroky AN, et al. Biases arising from linked administrative data for epidemiological research: a conceptual framework from registration to analyses. Eur J Epidemiol. 2022; 10.1007/s10654-022-00934-w.

40. Pattaro, S, Bailey, N, Dibben, C. Occupational differences in COVID-19 hospital admission and mortality between women and men in Scotland: a population-based study using inked administrative data. Int J Popul Data Sci. 2022; 10.23889/ijpds.v7i3.1841.

